# Structural Equation Modelling of Healthcare Quality, Patient Satisfaction, and Patient Loyalty in Health Insurance Hospitals in Alexandria, Egypt

**DOI:** 10.64898/2026.01.05.26343433

**Authors:** Doaa S. Galal, Magda R. Ahmed, Ekram W. Abd El-Wahab, Basem F. Abdel-Aziz

## Abstract

**Background:** Improving healthcare quality is fundamental to building patient trust, strengthening continuity of care, and improving service delivery. Within Egypt’s Health Insurance Organization (HIO), patient satisfaction and loyalty serve as critical performance indicators, particularly as the country transitions toward Universal Health Insurance (UHI).

**Methods:** A cross-sectional analytical study was conducted among 983 HIO beneficiaries from four hospitals in Alexandria. A validated questionnaire measured eight domains of healthcare quality along with patient satisfaction and loyalty. Construct validity was assessed through confirmatory factor analysis (CFA), and structural equation modelling (SEM) was used to examine the direct effects of healthcare quality on satisfaction and loyalty.

**Results:** CFA demonstrated strong model fit (CFI = 0.943; TLI = 0.940; RMSEA = 0.031), with all constructs showing high reliability (Cronbach’s α > 0.70). Healthcare quality had a substantial direct effect on patient satisfaction (β = 0.607) and a modest but significant direct effect on patient loyalty (β = 0.263). Satisfaction, likewise, exerted a strong direct influence on loyalty (β = 0.545), with all pathways reaching statistical significance.

**Conclusions:** Healthcare quality is a pivotal determinant of patient satisfaction and loyalty within HIO hospitals. Targeted improvements, particularly in communication, staff competence, responsiveness, and transitional care, can enhance patient-centered outcomes and support the successful implementation of Egypt’s UHI reforms.

**What is Already Known on This Topic:** Patient satisfaction and loyalty are widely used indicators of healthcare performance.

Perceived healthcare quality consistently predicts satisfaction and loyalty across health systems.

In Egypt—and particularly in Health Insurance Organization (HIO) hospitals—few studies have used structural equation modelling (SEM) to examine these relationships.

**What This Study Adds:** Delivers the first SEM-based assessment of healthcare quality, satisfaction, and loyalty among HIO beneficiaries in Alexandria.

Shows that perceived healthcare quality strongly predicts satisfaction and directly enhances loyalty.

Provides validated measurement models for eight quality domains within the Egyptian public insurance context.

How This Study Might Affect Research, Practice, or Policy

Identifies priority areas for quality improvement: communication, competence, responsiveness, and transition of care.

Supports Universal Health Insurance (UHI) reforms by highlighting predictors of trust and loyalty in public services.

Offers a validated framework for routine patient-experience monitoring across HIO hospitals.

## INTRODUCTION

Good health is a core component of societal well-being, and the World Health Organization (WHO) defines health as “a state of complete physical, psychological, and social well-being and not merely the absence of disease,” affirming that all individuals have the right to equitable healthcare services (1). Universal Health Insurance (UHI) has therefore become a global priority, ensuring access to preventive, curative, rehabilitative, and palliative healthcare without financial hardship (2). In Egypt, the right to free healthcare services is supported by national legislation (3), and patient satisfaction has long been recognized as a key indicator of healthcare system performance (4).

Egypt’s healthcare system is pluralistic, comprising governmental, public, and non-governmental sectors, including the Ministry of Health and Population (MOHP), the Health Insurance Organization (HIO), military hospitals, and private providers (5). Since its establishment in 1964, the HIO has continuously expanded its coverage, from government employees to students, infants, and newborns, reaching nearly 30 million beneficiaries by 2002 (6). Despite this expansion, considerable challenges persist: as of 2015, the HIO covered only 58% of Egyptians, and 32% remained unable to access needed healthcare services (7, 8). Out-of-pocket (OOP) payments accounted for nearly 60% of total health expenditure (7), reflecting persistent financial barriers.

Recent health sector reforms, including the launch of the National Health Insurance (NHI) system in 2018, aim to expand coverage, improve efficiency, and enhance service quality in alignment with Egypt’s Vision 2030 (9, 10). Improving healthcare quality is central to these reforms, as quality directly influences service effectiveness, safety, and patient trust (11, 12). Patient-centered healthcare emphasizes meeting patient needs and minimizing gaps between expectations and actual experiences (13–15).

Accordingly, patient satisfaction has emerged as a critical indicator of service quality, reflecting patients’ judgments of the care received (16, 17).

Patient loyalty, reflected in patients’ intention to reuse services and recommend them to others, is another important measure of healthcare performance (18). Loyalty is closely linked to perceived quality and satisfaction and plays a strategic role in strengthening trust, adherence, and long-term engagement with public healthcare services.

Structural Equation Modelling (SEM) provides a robust analytical framework for assessing such complex relationships. SEM is widely used in healthcare and social sciences to evaluate latent constructs and test causal pathways (14, 19). Previous studies indicate strong interrelationships between healthcare quality, patient satisfaction, and patient loyalty (14, 20, 21). However, in Egypt, empirical studies using SEM to examine these relationships within the public insurance system remain limited (22).

Given Egypt’s transition toward universal health coverage, understanding the determinants of satisfaction and loyalty among HIO beneficiaries is essential for improving service quality and ensuring successful implementation of the NHI. This study therefore employs SEM to examine how healthcare quality influences patient satisfaction and loyalty in Health Insurance hospitals in Alexandria, and to explore the mediating pathways among these constructs.

## METHODS

### Study Design, Setting and Population

The operational phase of the study extended from June 2022 to December 2023. During this period, data collection and fieldwork activities were carried out across the selected Health Insurance Organization (HIO) hospitals in Alexandria [Gamal Abdel Naser Hospital, Talba Sporting Hospital, Abou Kir Hospital, and Karmoz Hospital], where patients were interviewed across multiple settings including inpatient wards, outpatient clinics, emergency departments, and other service units. An analytical cross-sectional study design was employed. The target population consisted of patients attending these hospitals between September 2022 and December 2023. Eligible participants included HI beneficiary patients as well as parents of paediatric patients in Talaba Sporting Hospital, whereas individuals who declined participation were excluded.

### Sampling Design and Technique

A simple stratified random sampling technique was used to select HIO patients. Stratification was based on the four participating hospitals, and data were gathered from patients admitted to inpatient departments or attending outpatient, emergency, or other services, such as the renal dialysis unit. The number of participants was distributed equally among the hospitals, and data collection continued until the required sample size was reached. The sample size was determined according to the rule of thumb for SEM, which recommends at least ten cases per parameter. Given that the maximum anticipated number of parameters was eighty-seven, a minimum of 870 participants was required; this number was rounded to 900 participants (23). Ultimately, 983 beneficiaries were included, satisfying SEM requirements for latent constructs and model estimation.

### Data Collection Methods and Tools

Patients were interviewed face-to-face using a structured questionnaire composed of two main sections (File S1). The first section collected sociodemographic information such as age, sex, marital status, working status, residence, family size, dependents covered by insurance, occupation, education, and income. It also addressed current health problems, whether chronic or acute, as well as health insurance utilization, including hospital name, department visited, duration of service use, and frequency of visits.

The second section consisted of the structured Arabic version of the validated National Patient Satisfaction Survey (NPSS), a modified SERVQUAL-based tool widely used in studies conducted in the United Arab Emirates (20, 24). The NPSS includes eighty-seven items categorized into three core components: healthcare quality, patient satisfaction, and patient loyalty. Previous studies reported a Cronbach’s alpha of 0.961 for the full scale, demonstrating excellent internal reliability (25). Respondents rated each item on a five-point Likert scale ranging from “strongly disagree” (1) to “strongly agree” (5).

A pilot study of fifteen participants was conducted to pretest the Arabic questionnaire. The wording was found to be appropriate, and internal consistency was assessed using Cronbach’s alpha. The healthcare quality scale produced a reliability coefficient of 0.924, patient satisfaction 0.821, and patient loyalty 0.731, all exceeding the acceptable threshold of 0.7.

### Assessment of Healthcare Service Quality

The evaluation of healthcare quality components encompassed eight major domains. The first domain, tangibles and facilities, included nine items assessing tangible and physical aspects of service delivery along with four items related to the quality of ancillary services and facilities, resulting in a total of thirteen items (Figure S1). Courtesy and empathy constituted the second domain, containing seven items that focused on the interpersonal aspects of care (Figure S2). The third domain, responsiveness and physiological aspects, consisted of two items addressing responsiveness and psychosocial elements and six items concerning patient involvement, yielding eight items overall (Figure S3). The fourth domain evaluated competency, fairness, and trust, combining six items on fairness and trust with five items related to competency and confidence, for a total of eleven items (Figure S4). The fifth domain, information and communication, merged three items assessing communication with four items on the information provided to patients, producing seven items in total (Figure S5). The sixth domain covered availability, accessibility, and timeliness of services and included four items measuring timeliness, five items on waiting times and delays, and six items evaluating availability and accessibility, amounting to fifteen items (Figure S6). The seventh domain focused on the transition to home, incorporating seven items related to the discharge process and patients’ transition after hospitalization (Figure S7). Finally, the eighth domain addressed organization management, regulation, and payment, containing eight items related to organizational and regulatory aspects and three items concerning payment matters when applicable, for a total of eleven items (Figure S8).

### Evaluation of Healthcare Quality Outcomes

We measured healthcare quality outcomes through patient satisfaction and loyalty. Satisfaction items assessed perceptions of treatment effectiveness, administrative procedures, and ancillary services, as well as overall impressions of the hospital (26) (Figure S9).

Loyalty items evaluated willingness to return to the hospital, preference for using services even when alternatives exist, and readiness to recommend the services to others, including friends, colleagues, and family (27) (Figures S10-S13).

### Statistical analysis

Descriptive statistics such as frequencies, percentages, means, and standard deviations to summarize sample characteristics and scale scores. Confirmatory factor analysis (CFA) was conducted to assess the measurement properties of the healthcare quality, satisfaction, and loyalty constructs. Model fit was evaluated using multiple indices including χ²/df, Comparative Fit Index (CFI), Tucker–Lewis Index (TLI), Normed Fit Index (NFI), Incremental Fit Index (IFI), Root Mean Square Error of Approximation (RMSEA), and Standardized Root Mean Square Residual (SRMR). Structural equation modelling (SEM) was subsequently applied to examine the direct and indirect relationships among healthcare quality, patient satisfaction, and patient loyalty.

## RESULTS

### Characteristics of the study population

Most respondents were female (57.6%) and lived in urban areas (73.1%). The mean age was 45.1 ± 11.05 years, with the majority clustered between 30 and 50 years (27.7% aged 30–40 and 34% aged 40–50). Nearly half (52.4%) were employed. Educational attainment was mainly secondary or intermediate (48.1%), and income inadequacy was reported by 42.8% of participants. Most were married (67.2%), with typical family sizes of three to four members (50.3%). Regarding insurance coverage, 57.6% reported coverage for one to two family members, while 34.8% reported three to four members (Table 1). About 54.6% of respondents reported chronic health conditions, while 45.4% experienced acute issues. Nearly half (48.4%) had been using health insurance services for over six years; 31.4% reported only two to three service encounters (Table S1).

**Table 1.**
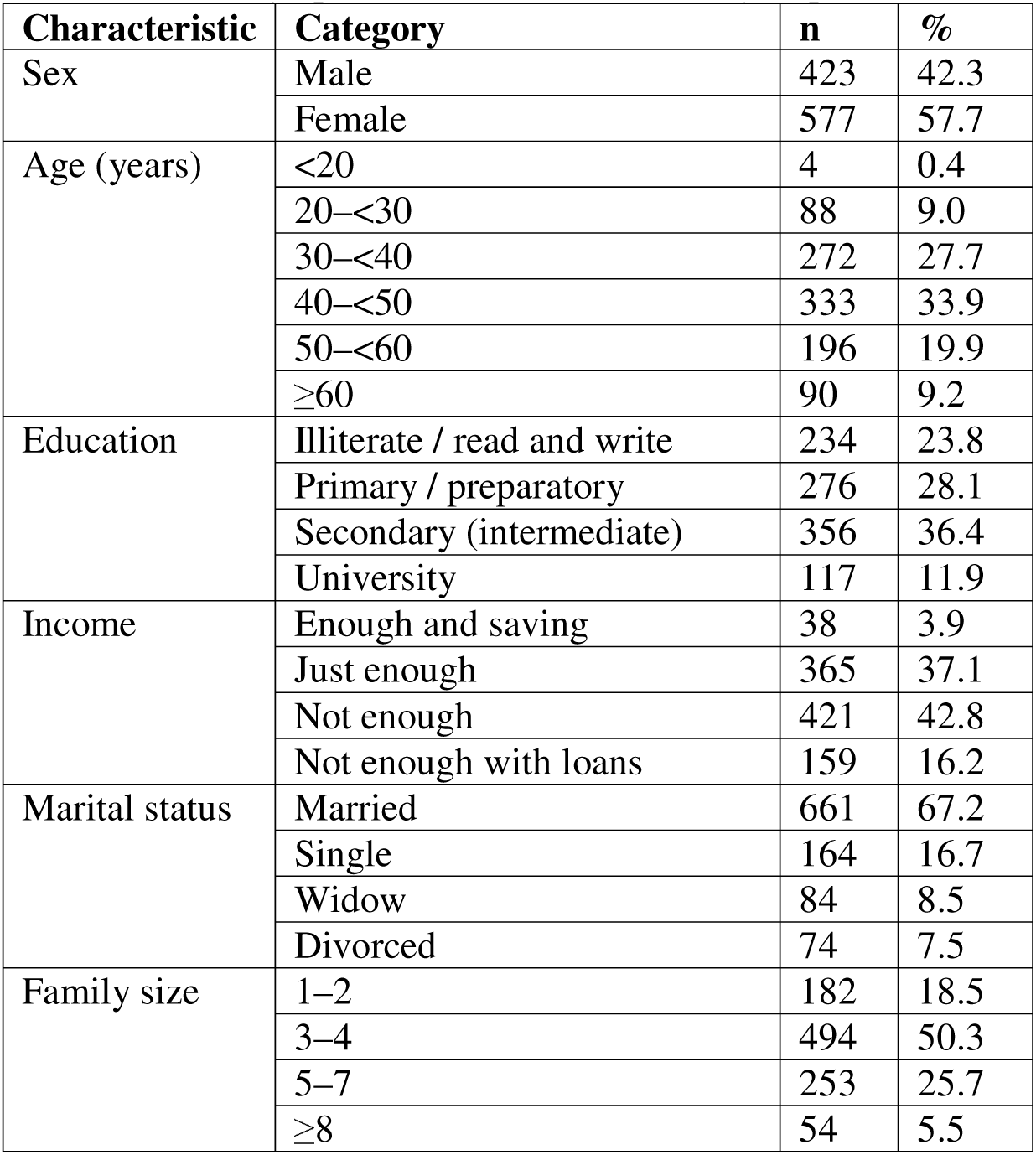
Sociodemographic Characteristics of Study Population

### Overall Quality, Satisfaction, and Loyalty

Most participants (61.7%) rated overall service quality as moderate, while only 27.1% described it as good or very good, highlighting considerable room for improvement. In contrast, patient satisfaction displayed a more positive distribution: nearly half of respondents (49.8%) reported good or very good satisfaction, although 23.4% indicated low or very low satisfaction. Patient loyalty showed an even stronger pattern, with 40.1% rating their loyalty as good or very good and only 24.9% reporting low or very low loyalty. Collectively, these results suggest that while perceptions of core service quality remain modest, satisfaction and loyalty are comparatively more robust, implying that targeted enhancements in specific quality domains may further strengthen overall patient engagement and retention (Table 2).

**Table 2.** Evaluation of Overall Quality, Patient Satisfaction, and Patient Loyalty

| **Level** | **Overall Quality** | **Patient Satisfaction** | **Patient Loyalty** |
| --- | --- | --- | --- |
|  | **n (%)** | **n** | **n (%)** |
| Very low | 13 (1.3) | 72 | 105 (10.7) |
| Low | 96 (9.8) | 162 | 140 (14.2) |
| Moderate | 607 (61.7) | 337 | 344 (35.0) |
| Good | 198 (20.1) | 215 | 173 (17.6) |
| Very good | 69 (7.0) | 197 | 221 (22.5) |

### Reliability of Study Constructs

Table 3 summarizes descriptive statistics and reliability of the endogenous and exogenous Variables using Cronbach’s alpha. All scales exhibited high internal consistency, with alpha values exceeding 0.80. The “Overall Health Care Quality” scale achieved the highest reliability (α = 0.937), while the “Patient Loyalty” scale, although lower (α = 0.769), still met the acceptable threshold. Among the measured dimensions, “Information and Communication” demonstrated the highest average response (3.291 ± 0.063), followed by “Availability of Resources” (3.264 ± 0.084) and “Patient Loyalty” (3.258 ± 0.032).

**Table 3.**
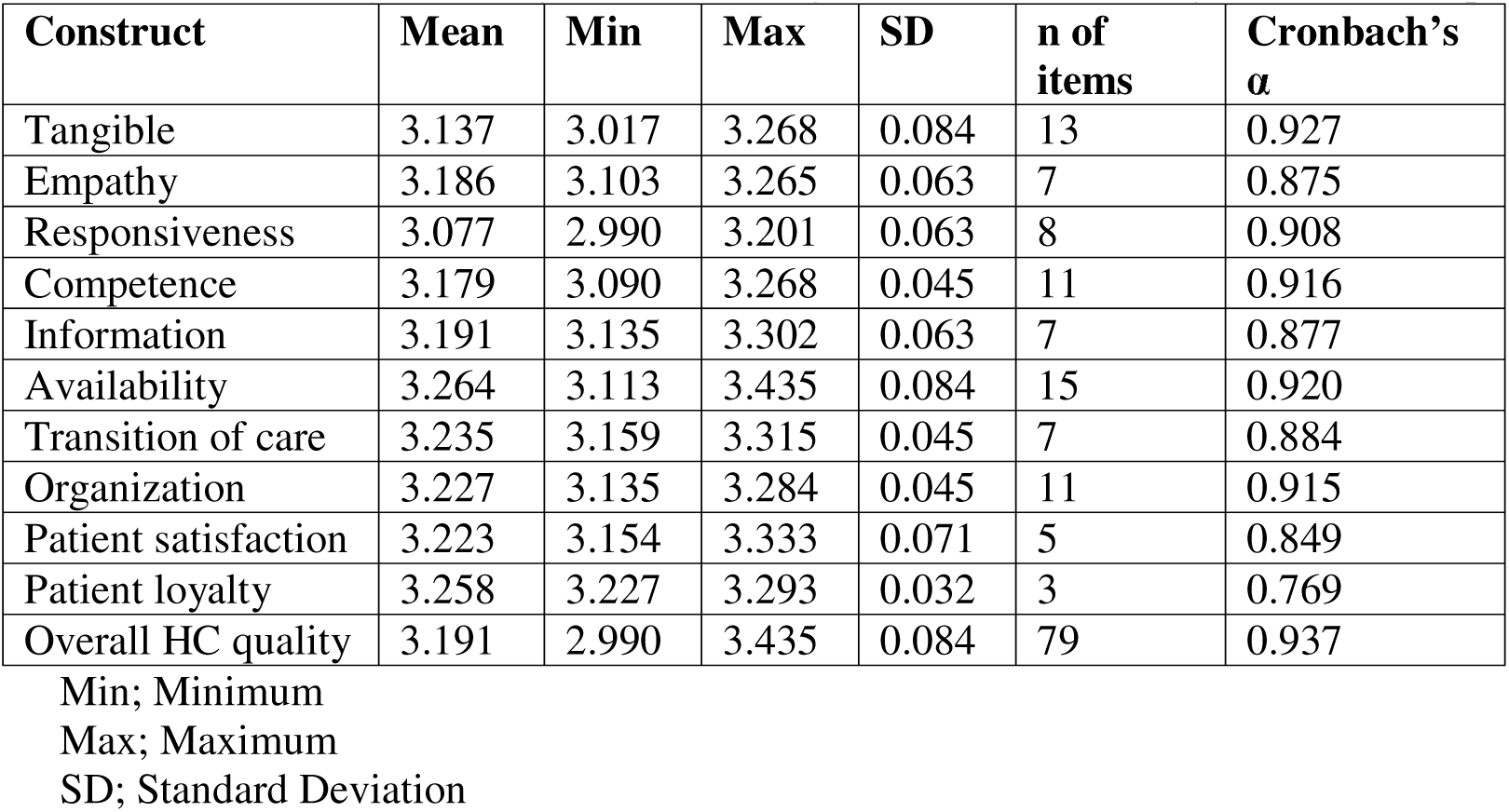
Reliability of Endogenous and Exogenous Variables Using Cronbach’s Alpha

| **Construct** | **Mean** | **Min** | **Max** | **SD** | **n of items** | **Cronbach's $\alpha$** |
| --- | --- | --- | --- | --- | --- | --- |
| Tangible | 3.137 | 3.017 | 3.268 | 0.084 | 13 | 0.927 |
| Empathy | 3.186 | 3.103 | 3.265 | 0.063 | 7 | 0.875 |
| Responsiveness | 3.077 | 2.990 | 3.201 | 0.063 | 8 | 0.908 |
| Competence | 3.179 | 3.090 | 3.268 | 0.045 | 11 | 0.916 |
| Information | 3.191 | 3.135 | 3.302 | 0.063 | 7 | 0.877 |
| Availability | 3.264 | 3.113 | 3.435 | 0.084 | 15 | 0.920 |
| Transition of care | 3.235 | 3.159 | 3.315 | 0.045 | 7 | 0.884 |
| Organization | 3.227 | 3.135 | 3.284 | 0.045 | 11 | 0.915 |
| Patient satisfaction | 3.223 | 3.154 | 3.333 | 0.071 | 5 | 0.849 |
| Patient loyalty | 3.258 | 3.227 | 3.293 | 0.032 | 3 | 0.769 |
| Overall HC quality | 3.191 | 2.990 | 3.435 | 0.084 | 79 | 0.937 |

Min; Minimum
Max; Maximum
SD; Standard Deviation

### Measurement Model Evaluation: Assessment of Health Care Quality Domains (CFA)

Across the nine domains, confirmatory factor analyses (CFAs) showed solid factor loadings (FLs) and high reliability. Tangibles, Empathy, Responsiveness, Competence, Information, Availability, Organization, Transition of Care, and Patient Satisfaction all demonstrated FLs generally between 0.65 and 0.77 and Cronbach’s α > 0.80, with good-to-excellent model fit (Figures S14–S22). The Patient Loyalty scale also showed acceptable reliability (α = 0.769) (Figure S23).

### Conceptual Model and Path Analysis: Health Care Quality, Satisfaction, and Loyalty

The conceptual model specified a direct relationship between healthcare quality, patient satisfaction, and patient loyalty (Figure 1). Path Analysis of Health Care Quality, Patient Satisfaction, and Patient Loyalty revealed that Health Care quality strongly predicts patient satisfaction (β = 0.607) and moderately predicts loyalty (β = 0.263), while satisfaction substantially drives loyalty (β = 0.545). Model fit indices (e.g., GFI = 0.977, CFI = 0.998, RMSEA = 0.059) confirmed model adequacy (Figure S24).

**Figure 1.**
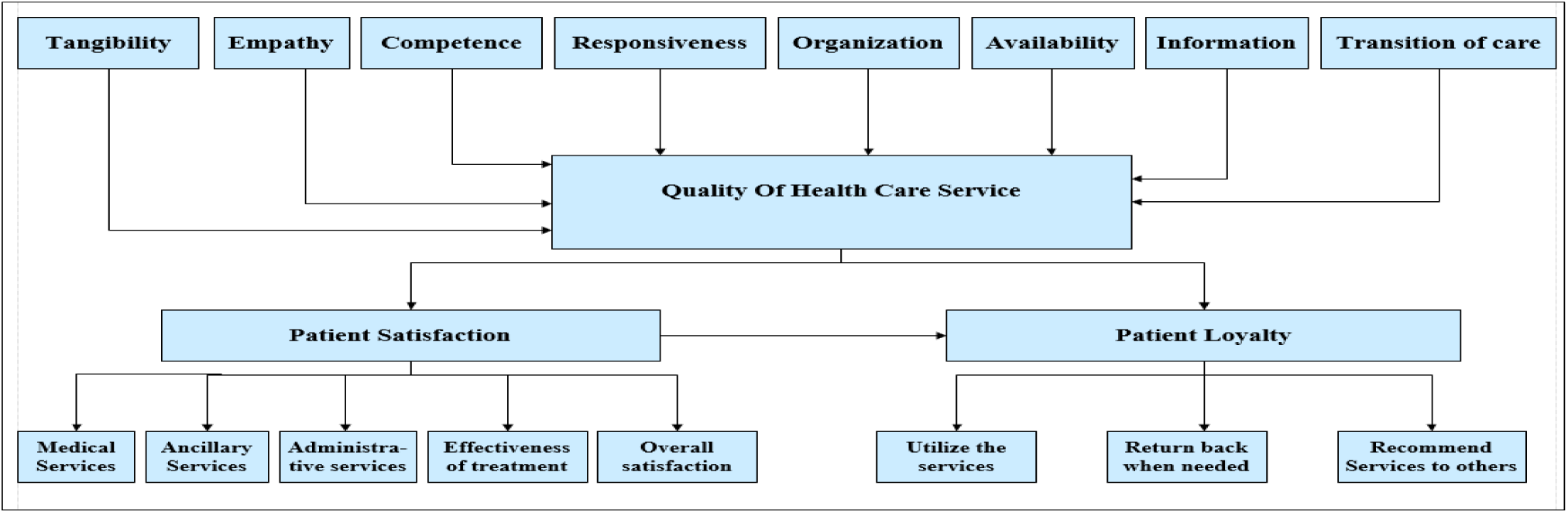
Conceptual model of the relationships between Health Care quality, patient satisfaction, and patient loyalty.

### CFA, Reliability, and Validity

A total of 17 latent variables were excluded from the overall model [consisting of 79 latent variables] due to low FLs (< 0.7) (Table S2).

### Initial and Modified Measurement Model of Health Care Quality

An initial healthcare quality model (Figure S25), demonstrating acceptable fit (CMIN/DF = 1.926; RMSEA = 0.032). Following modifications, the model showed improved model indices (CMIN/DF = 1.851; RMSEA = 0.031) (Figure S26). All FLs in Table S3 were significant and greater than 0.7, confirming construct validity.

### Reliability and validity measures

Composite reliability ranged from 0.849 to 0.919, while AVE values (0.502–0.553) confirmed convergent validity. Discriminant validity was achieved as AVE square roots exceeded inter-factor correlations. The maximum reliability (MaxR(H)) values were all above 0.85 (Table 4).

**Table 4.** Reliability and Validity of Healthcare Quality Domains

| Domain/<br>Constructs | $\alpha$ | CR | AVE | MSV | Max R (H) | Transition | Tangible | Information | Response | Competent | Available | Organize | Empathy |
| --- | --- | --- | --- | --- | --- | --- | --- | --- | --- | --- | --- | --- | --- |
| Transition | 0.884 | 0.884 | 0.521 | 0.241 | 0.885 | 0.72 |  |  |  |  |  |  |  |
| Tangible | 0.927 | 0.910 | 0.502 | 0.084 | 0.910 | 0.29 | 0.71 |  |  |  |  |  |  |
| Information | 0.877 | 0.849 | 0.529 | 0.215 | 0.849 | 0.36 | 0.27 | 0.73 |  |  |  |  |  |
| Response | 0.908 | 0.908 | 0.553 | 0.085 | 0.909 | 0.20 | 0.07 | 0.23 | 0.74 |  |  |  |  |
| Competent | 0.916 | 0.882 | 0.518 | 0.115 | 0.883 | 0.34 | 0.29 | 0.31 | 0.09 | 0.72 |  |  |  |
| Available | 0.920 | 0.919 | 0.507 | 0.100 | 0.919 | 0.21 | 0.1 | 0.24 | 0.29 | 0.18 | 0.71 |  |  |
| Organize | 0.915 | 0.893 | 0.511 | 0.089 | 0.894 | 0.17 | 0.06 | 0.3 | -0.02 | 0.08 | 0.08 | 0.72 |  |
| Empathy | 0.875 | 0.861 | 0.507 | 0.241 | 0.862 | 0.49 | 0.2 | 0.46 | 0.21 | 0.19 | 0.32 | 0.22 | 0.71 |

$\alpha$ :Cronbach's alpha
CR; composite reliability
AVE; Average Variance Extracted
MSV; Maximum Shared Variance
MaxR(H); Maximum H Reliability

### Combined Measurement Model for Health Care Quality, Patient Satisfaction, and Loyalty

The initial combined CFA model (Figure S27) showed acceptable fit (CMIN/DF = 1.923; RMSEA = 0.032). After applying modification indices (Figure 2), the model improved further (CMIN/DF = 1.905).

**Figure 2:**
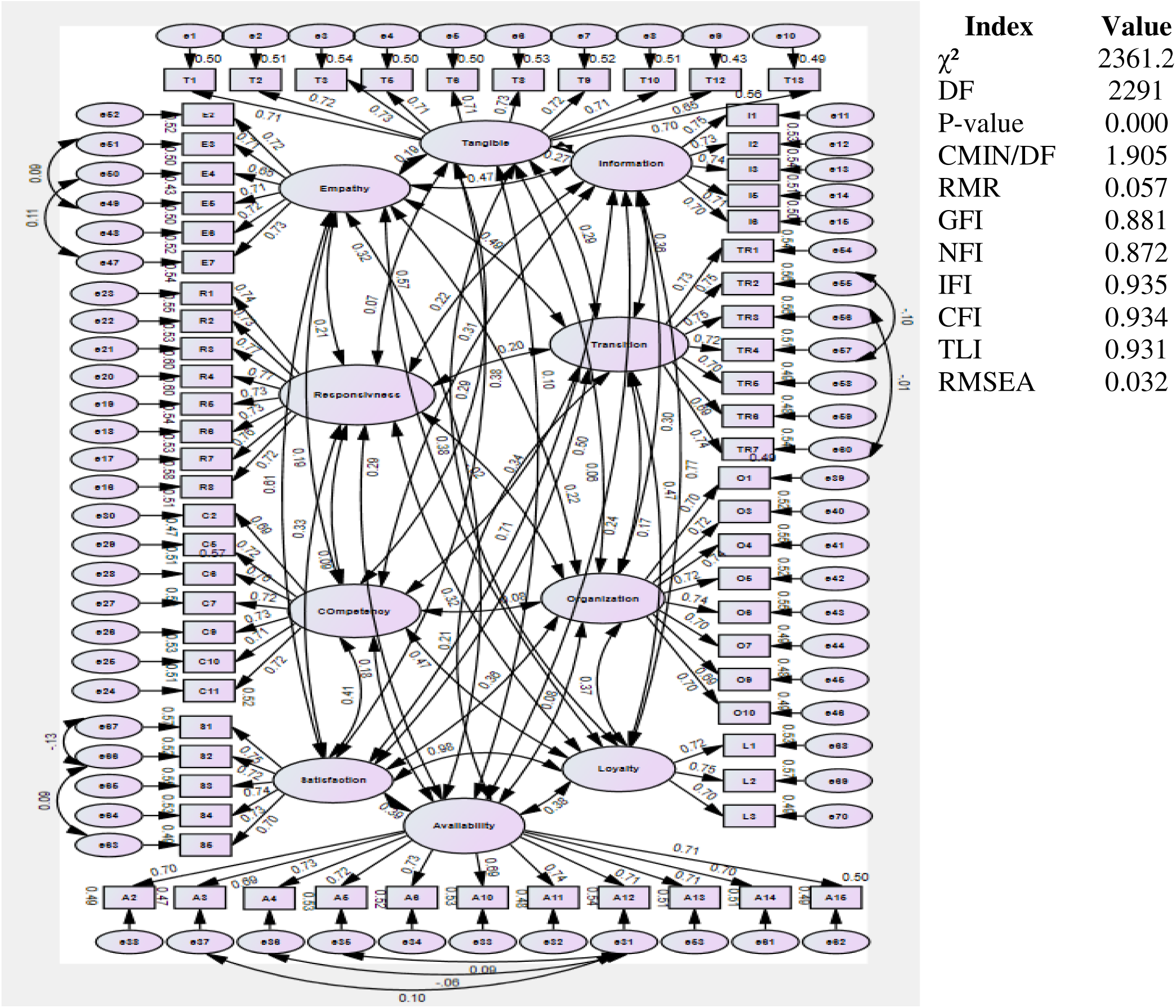
Modified health care quality, patient satisfaction, and patient loyalty constructs

### FL and regression of Health Care Quality, Patient Satisfaction, and Patient Loyalty Scale

All standardized FLs exceeded 0.7 and were statistically significant (p < 0.05), with standardized loadings ranging from 0.654 to 0.774, confirming the strong contribution of these items to the healthcare quality model (Table S4).

### Reliability and Construct Validity Measures for Health Care Quality, Patient Satisfaction, and Patient Loyalty

All constructs demonstrated strong reliability, with composite reliability values exceeding 0.7 (0.769–0.927), consistent with Cronbach’s alpha results. Convergent validity was supported by AVE values ranging from 0.501 to 0.544, all above the 0.50 threshold. The square roots of the AVE values (0.708–0.744) exceeded inter-construct correlations, and MSV values were lower than corresponding AVEs for all constructs, confirming adequate discriminant validity. Maximum R(H) values (0.772–0.919) further supported the robustness of the measurement model (Table 4).

### Structural Equation Modeling (SEM)

Figure S28 presented a well-fitting structure model (CMIN/DF = 1.975; RMSEA = 0.033) of the relationship between healthcare quality and the observed variables. Satisfaction and loyalty maintained acceptable fit (Figure S29). Following modifications (Figure 3), the model fit improved considerably. The initial pooled model (Figure S30) showed moderate fit, but after refinement (Figure S31), fit indices improved substantially (CFI = 0.973; RMSEA = 0.042).

**Figure 3:**
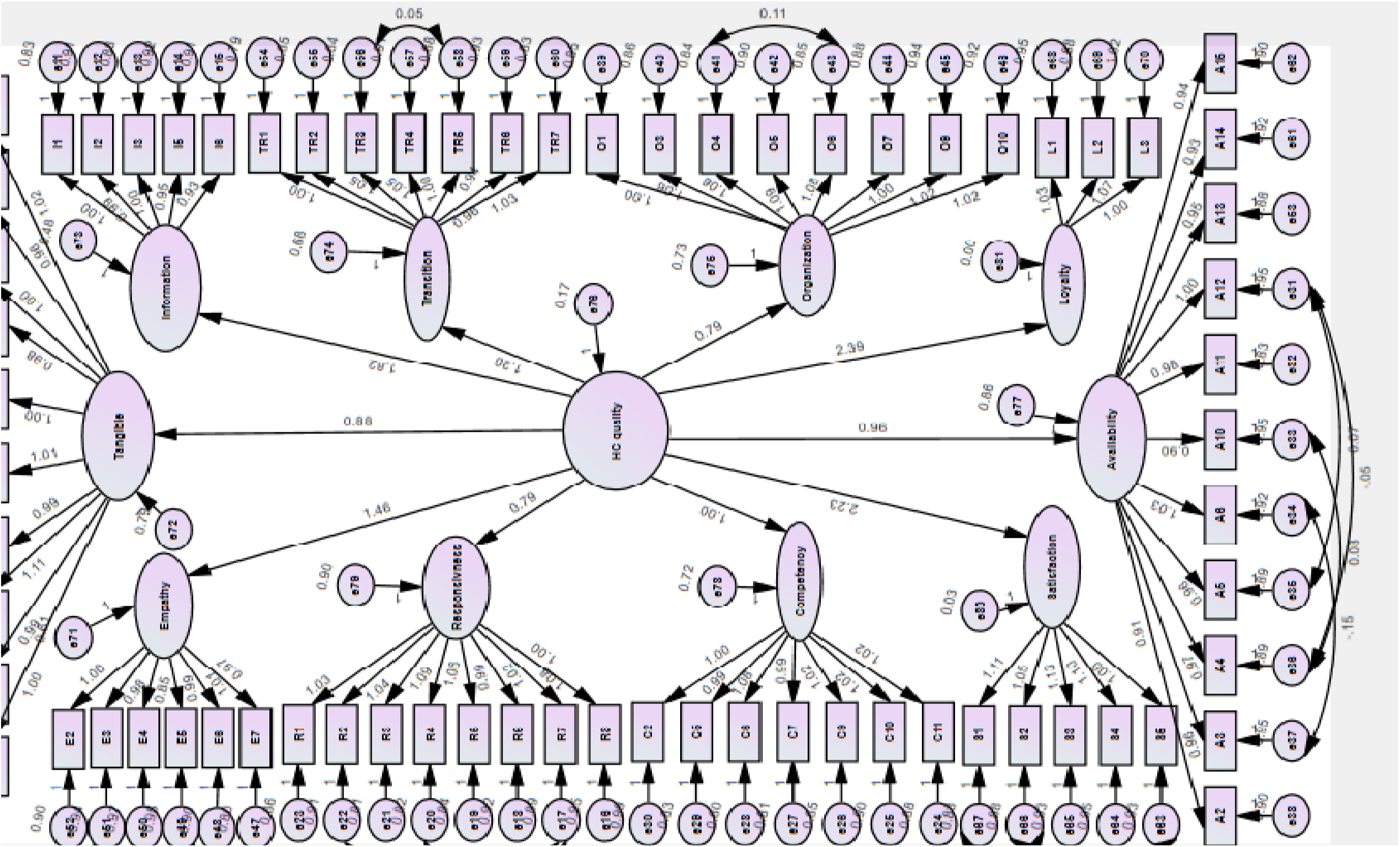
The modified relationship between Health Care quality, patient satisfaction, and loyalty observed variables

All paths from healthcare quality to its eight observed dimensions were significant, with the strongest effects for Transition (β = 0.632) and Empathy (β = 0.622), and moderate effects for Tangibles (β = 0.476) and Responsiveness (β = 0.276). Thus, the null hypothesis (H1) was rejected.

### Direct Causal Effects among Health Care Quality, Satisfaction, and Loyalty

The structural model in Figure 4 confirmed that healthcare quality significantly predicted both satisfaction and loyalty. Overall model fit was strong (CFI = 0.967; RMSEA = 0.043), supporting H2. Table S5 reported significant FLs (0.319–0.743), thus reinforcing construct validity.

**Figure 4:**
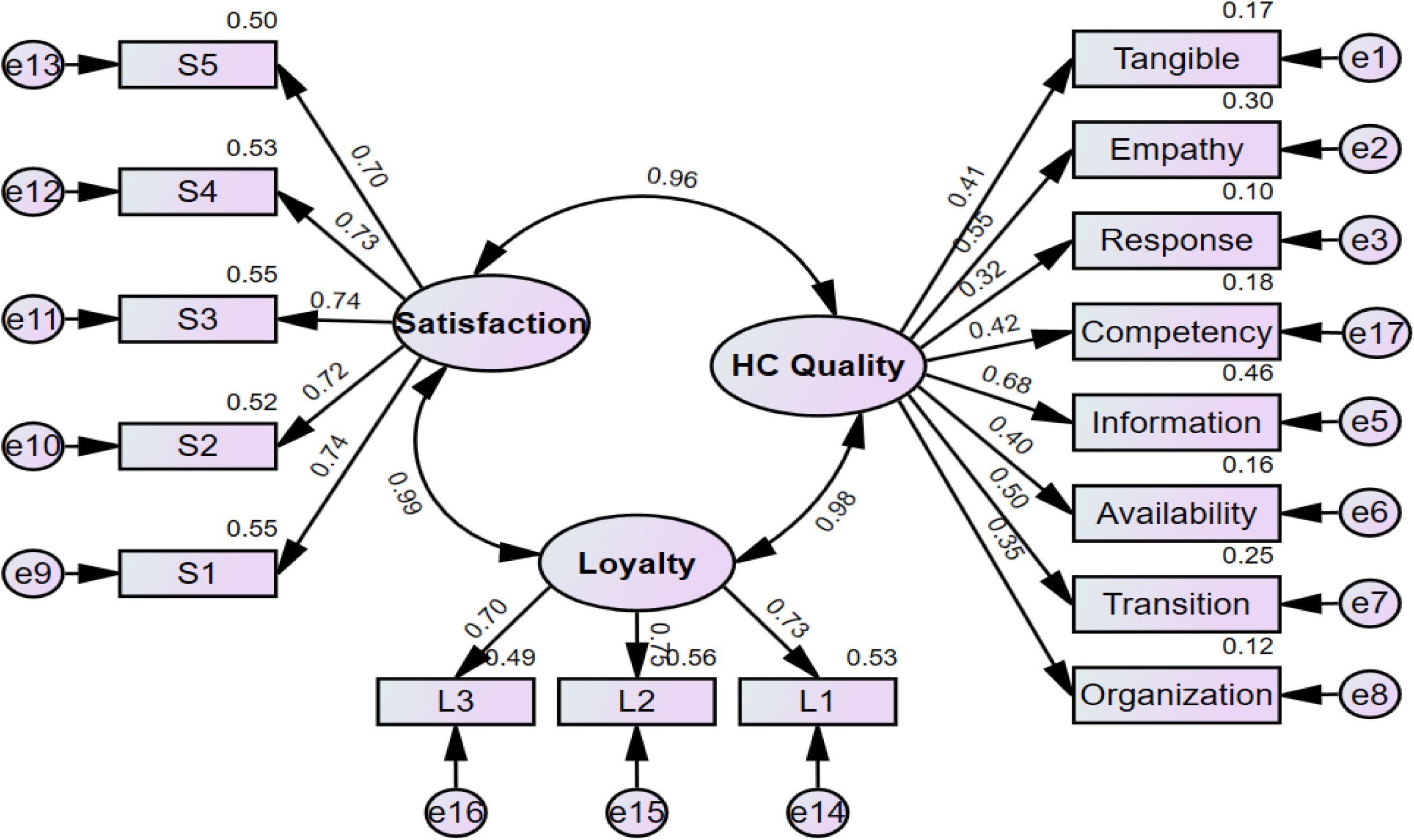
Causal Relationship Between Health Care Quality, Patient Satisfaction, and Patient Loyalty

### Total Effects of Health Care Quality on Satisfaction and Loyalty

Both satisfaction and loyalty were significantly influenced by healthcare quality (Figure 5). A one-SD increase in healthcare quality led to a 1.001 SD increase in satisfaction and a 0.982 SD increase in loyalty. Fit indices indicated excellent model alignment (CFI = 0.966; RMSEA = 0.043).

**Figure 5.**
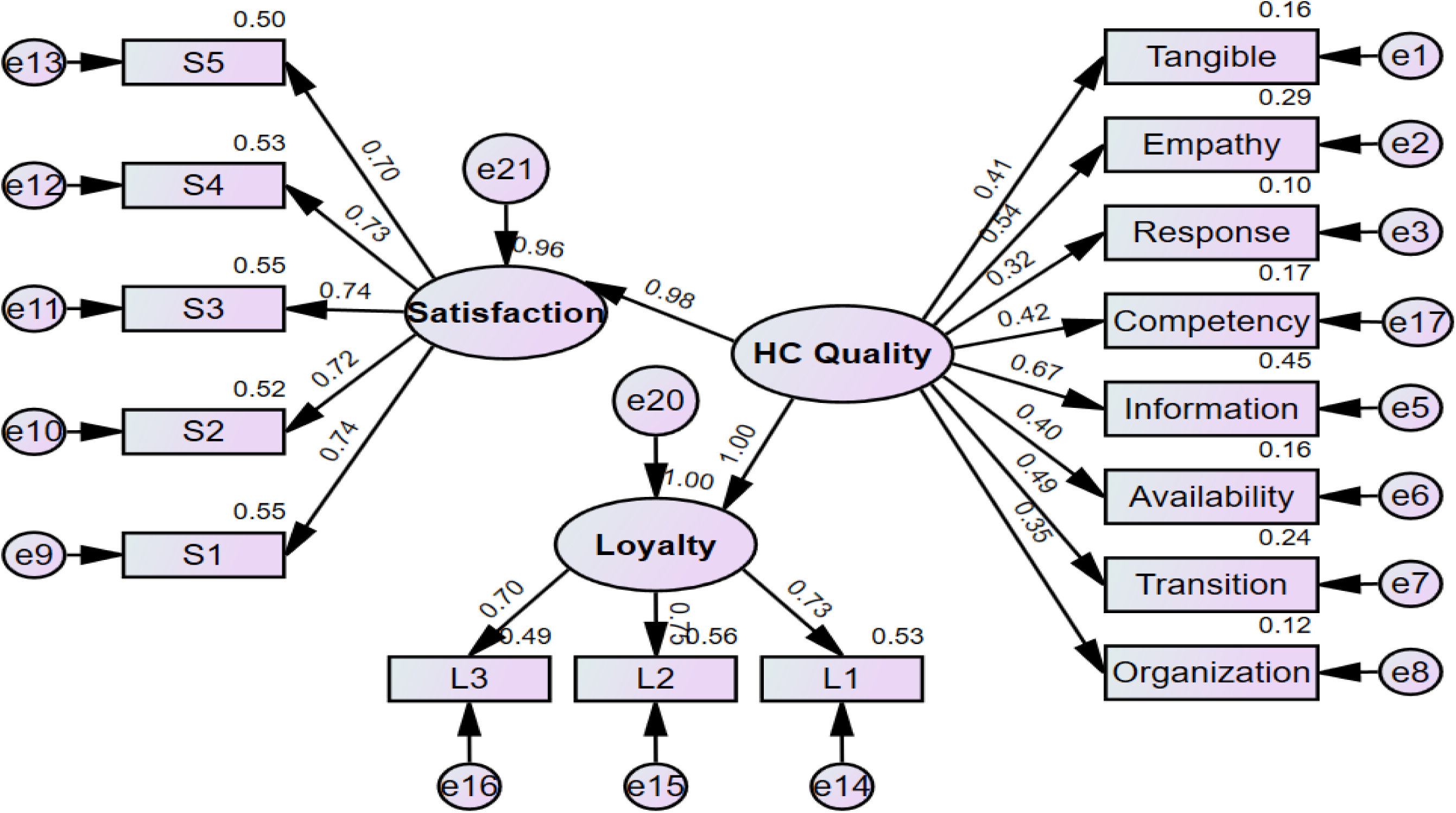
: Equal effect of Health Care quality on patient satisfaction and patient loyalty

### Two-Path Model: Health Care Quality → Satisfaction → Loyalty

Figure S32 tested the indirect pathway. healthcare quality did not significantly influence loyalty directly (p = 0.119), leading to acceptance of the null hypothesis (H4.1). However, healthcare quality significantly influenced satisfaction (p = 0.012), which in turn positively predicted loyalty (H4.2 supported). Model fit remained excellent (CFI = 0.967; RMSEA = 0.043).

## DISCUSSION

This study provides compelling evidence that healthcare quality strongly and directly shapes patient satisfaction and loyalty among HIO beneficiaries, reinforcing extensive international literature that identifies communication, competence, responsiveness, and organizational systems as core determinants of patient experience (28, 29). By integrating eight interrelated domains, tangibles, empathy, responsiveness, competency, information and communication, availability of services, organizational processes, and transition of care, the validated measurement model demonstrates that healthcare quality is a multidimensional construct that extends beyond traditional SERVQUAL indicators. This aligns with global findings emphasizing that patient experiences are shaped not only by interpersonal interactions but also by system-level processes that support continuity, coordination, and accessibility of care.

The results echo earlier studies showing that dimensions such as tangibles, responsiveness, communication, and accessibility have consistent impacts on patient satisfaction across different settings (30–37). These findings emphasize that patients evaluate healthcare not only based on clinical interactions but also through physical infrastructure, clarity of information, waiting time, organizational efficiency, and the attitudes and professionalism of staff. The current study’s structural models confirm these relationships, showing that improvements in healthcare quality consistently led to higher patient satisfaction, reinforcing this relationship across various contexts.

Crucially, the study highlights that satisfaction plays a central mediating role between healthcare quality and patient loyalty. Although healthcare quality initially appears to influence loyalty directly, this direct effect becomes non-significant once satisfaction is accounted for, indicating that patients commit to a healthcare provider primarily because they feel satisfied. not merely because quality is objectively high. This mirrors findings from prior research across health systems, which consistently show that satisfaction strengthens or fully mediates the relationship between perceived quality and loyalty (38–44).

This pattern has important implications for public insurance contexts. Unlike private health systems, where loyalty may reflect competitive consumer choice, loyalty in public insurance structures is vital for sustaining system utilization, ensuring continuity of care, reducing unnecessary reliance on the private sector, and maintaining the financial viability of public health reforms. The strong effect of satisfaction on loyalty underscores the need for patient-centred interventions, such as enhancing communication practices, strengthening care coordination, improving staff engagement, and optimizing facility organization, to build trust, adherence, repeated service use, and positive recommendations within communities.

The findings also support international evidence showing that patient satisfaction is shaped not only by clinical outcomes but by the total experience of care, including interactions with staff, clarity of communication, facility conditions, wait times, and fairness of administrative processes (45–47). As such, healthcare quality improvement strategies must be comprehensive, targeting both clinical and non-clinical aspects of care delivery.

Overall, the eight-domain healthcare quality model validated in this study offers a robust, standardized assessment tool capable of guiding quality monitoring and improvement across HIO hospitals. Its strong psychometric performance and alignment with international evidence make it suitable for broader implementation as part of the expanding UHI system. By adopting such a holistic quality framework, policymakers and healthcare leaders can more effectively design patient-centred reforms, enhance satisfaction, and foster loyalty, ultimately promoting stronger engagement with public healthcare services and contributing to a more sustainable, trusted, and equitable health system.

The findings of this study highlight several important policy implications for strengthening healthcare quality within HIO hospitals. Improving communication and information-sharing between providers and patients is essential for enhancing understanding, transparency, and trust. Likewise, investing in staff competence through targeted training and continuous professional development can significantly elevate the overall standard of care. Responsiveness may be improved by optimizing workflow processes and reducing waiting times, which remain persistent concerns in public healthcare settings. Standardizing transitional care practices, including clear discharge instructions and structured follow-up plans, would further support continuity and patient safety. Integrating continuous patient-experience measurements into HIO monitoring systems is also critical for guiding quality improvement efforts, while evidence-based, patient-centred approaches can better support the broader goals of national UHI reform.

The findings have important implications for COVID-19–related clinical practices and policy development. The strong influence of healthcare quality on patient satisfaction, as well as its substantial indirect effect on loyalty, highlights the need for COVID care systems to prioritize improvements in communication, empathy, transition of care, and resource availability. Although overall perceived service quality was only moderate, relatively higher satisfaction and loyalty indicate that targeted enhancements in measurable quality domains can meaningfully strengthen patient engagement, adherence to treatment, and continuity of care during infectious disease crises. For policymakers, the validated measurement model offers a reliable framework for evaluating COVID-19 service performance and guiding quality-improvement initiatives, particularly in settings with strained resources. Strengthening patient-centered practices, ensuring smoother care transitions, and addressing disparities in access, particularly for populations reporting chronic conditions or financial constraints, can enhance trust in health systems and improve uptake of preventive and therapeutic COVID-19 services In conclusion, this study confirms that healthcare quality is a significant predictor of both patient satisfaction and loyalty in HIO hospitals. Enhancing key quality domains, particularly communication, staff competence, responsiveness, and transition of care, will strengthen patient trust, improve continuity of care, and contribute to the successful implementation of Egypt’s UHI goals. These findings underscore the importance of sustained investment in quality improvement as a central component of health system transformation.

## Study Limitations

The study faced several challenges that should be acknowledged. The approval process from the Ministry of Health and the Health Insurance Organization was lengthy and complex, resulting in delays in initiating data collection. Additionally, many patients withdrew from the study after receiving care, often unwilling to wait to complete the interview, which contributed to participant’s attrition. Some respondents appeared hesitant or less transparent in their answers, likely influenced by the presence of healthcare providers. Finally, incomplete or missing responses required the replacement of participants to maintain sample integrity. Despite these challenges, the study achieved its objectives and provides valuable insights to inform healthcare quality improvement within the HIO and the evolving UHI system.

## Ethical consideration

### Ethical approval and consent to participate

The study was approved by the institutional review board and the ethics committee of the High Institute of Public Health affiliated with Alexandria University, Egypt. We sought the permission and support of the local health authorities to conduct the study in the selected districts in Alexandria. The study was conducted in accordance with the international ethical guidelines and of the Declaration of Helsinki. Informed written consent was obtained from each participant after explaining the aim and concerns of the study. Data sheets were coded by number to ensure anonymity and confidentiality of the participants’ data.

This article does not contain any studies with animals performed by any of the authors.

- **Informed consent:** Declared
- **Funding:** None
- **Conflicts of interest:** None to declare.

## Consent for publication

All authors approved the manuscript for publication

## Availability of supporting data

All data are fully available without restriction by the corresponding author

## Data Availability

All data produced in the present study are available upon reasonable request to the authors

## Acknowledgements

We would like to acknowledge the study participants for accepting to participate in the study.

