## Supplementary material for "Structural Equation Modelling of Healthcare Quality, Patient Satisfaction, and Patient Loyalty in Health Insurance Hospitals in Alexandria, Egypt": Suppl tables

Supplementary tables

Table S1: Health Information about the study subjects:

| **Characteristics** | **Number** | **%** |
| --- | --- | --- |
| **Health Problem:**  Chronic  Acute | 537  446 | 54.6  45.4 |
| **Duration of using Health Insurance services:**  From 1-2 years  From 3-4 years  From 5-6 years  More than 6 years | 91  172  244  493 | 9.3  17.5  24.8  48.4 |
| **Frequency of using the services:**  First time  From 2-3 times  From 4 to 5 times  6 times or more | 279  309  208  187 | 28.4  31.4  21.2  19.0 |

Table S2: List of deleted variables from the model

| **Domain** | **Code** | **Question number** | **FL** | **Number of items** | **Deleted items** |
| --- | --- | --- | --- | --- | --- |
| **Tangible** | T4 | The hospital had a comfortable rest area for patients | 0.68 | 13 | 3 |
|  | T7 | The materials associated with the services (towels, etc.) were visually appealing | 0.66 |  |  |
|  | T11 | Communication facilities were available for the patients (telephones) | 0.66 |  |  |
| **Empathy** | E1 | I feel that the staff (doctors, nurses, etc.) showed a sincere interest in my condition | 0.68 | 7 | 1 |
| **Competency** | C1 | I felt that one doctor said something, while another doctor said something quite different | 0.68 | 11 | 4 |
|  | C3 | I felt that some patients enjoyed first-class treatment while many others did not | 0.68 |  |  |
|  | C4 | I felt that the doctors were talking about me, as I was not there | 0.68 |  |  |
|  | C8 | I had full confidence and trust in the doctor treating me | 0.69 |  |  |
| **Information** | I4 | Adequate information about my condition or treatment was given to my family. | 0.66 | 7 | 2 |
|  | I7 | While I was in certain units, I got enough information about the medical treatment I needed | 0.69 |  |  |
| **Availability** | A1 | After checking in at the hospital, I had to wait before going to my room | 0.4 | 15 | 4 |
|  | A7 | My discharge was handled in a timely manner | 0.66 |  |  |
|  | A8 | The medical procedures were done correctly the first time | 0.69 |  |  |
|  | A9 | When I needed help, I got help in time | 0.69 |  |  |
| **Organization** | O2 | I feel that the rules of the hospital were always strictly maintained | 0.69 | 11 | 3 |
|  | O8 | I feel that the rules of the hospital were always strictly maintained | 0.69 |  |  |
|  | O11 | When I had to pay for the treatment, the overall cost was reasonable | 0.64 |  |  |

Table S3: Standardized and Unstandardized Regression Weights for Health Care Quality

| Code | Question | Relation | HC Quality Subscales | Unstandardized Regression Weights | | | | Standardized Regression  Weights |
| --- | --- | --- | --- | --- | --- | --- | --- | --- |
|  |  |  |  | **Estimate** | **S.E.** | **C.R.** | **P** | **Estimate** |
| Q_1 | The cleanliness throughout the hospital | <--- | Tangible | 1 |  |  |  | 0.709 |
| Q_2 | The cabins/wards were clean. | <--- | Tangible | 0.982 | 0.049 | 20.22 | *** | 0.718 |
| Q_3 | The doctors, nurses, and staff were neatly dressed. | <--- | Tangible | 1.094 | 0.053 | 20.766 | *** | 0.738 |
| Q_5 | The hospital has modern-looking equipment. | <--- | Tangible | 0.98 | 0.049 | 20 | *** | 0.71 |
| Q_6 | The hospital used state-of-the-art medical equipment. | <--- | Tangible | 0.978 | 0.049 | 20.025 | *** | 0.711 |
| Q_8 | The physical facilities were visually appealing. | <--- | Tangible | 1.012 | 0.049 | 20.667 | *** | 0.734 |
| Q_9 | The toilets were clean. | <--- | Tangible | 0.983 | 0.048 | 20.571 | *** | 0.731 |
| Q_10 | Adequate parking was available. | <--- | Tangible | 0.993 | 0.049 | 20.117 | *** | 0.714 |
| Q_12 | I found the taste of my food to be very good. | <--- | Tangible | 0.882 | 0.05 | 17.549 | *** | 0.623 |
| Q_13 | Specially assigned rooms were available. | <--- | Tangible | 0.971 | 0.051 | 18.918 | *** | 0.671 |
| Q_40 | It was easy for me to find someone to talk with. | <--- | Information | 1 |  |  |  | 0.738 |
| Q_41 | My family had enough opportunities to talk to my doctors. | <--- | Information | 1.004 | 0.05 | 20.237 | *** | 0.729 |
| Q_42 | My family had enough opportunities to talk to the nurses. | <--- | Information | 1.036 | 0.05 | 20.88 | *** | 0.754 |
| Q_44 | The hospital informed me about the medical options. | <--- | Information | 0.962 | 0.049 | 19.809 | *** | 0.713 |
| Q_45 | I got enough information about my medical condition. | <--- | Information | 0.931 | 0.048 | 19.454 | *** | 0.7 |
| Q_28 | I got answers that I could understand from doctors. | <--- | Response | 1 |  |  |  | 0.716 |
| Q_27 | I got answers that I could understand from the Nurse. | <--- | Response | 1.092 | 0.051 | 21.629 | *** | 0.76 |
| Q_26 | The nursing staff explained the treatment simply. | <--- | Response | 1.021 | 0.049 | 20.755 | *** | 0.729 |
| Q_25 | Someone explained to me the reasons why I had to wait. | <--- | Response | 0.99 | 0.048 | 20.82 | *** | 0.731 |
| Q_24 | I had said enough about the medical treatment. | <--- | Response | 1.082 | 0.049 | 22.027 | *** | 0.774 |
| Q_23 | The hospital involved me in the decision about my care. | <--- | Response | 1.088 | 0.05 | 21.93 | *** | 0.771 |
| Q_22 | When I had some anxiety or fear, the nurse discussed it. | <--- | Response | 1.036 | 0.05 | 20.809 | *** | 0.731 |
| Q_21 | When I had some anxiety or fear, the doctor discussed it. | <--- | Response | 1.025 | 0.049 | 21.08 | *** | 0.741 |
| Q_38 | The hospital did all it could to help control my pain. | <--- | Competency. | 1.027 | 0.054 | 19.123 | *** | 0.717 |
| Q_37 | I had full confidence and trust in the nurse treating me. | <--- | Competency. | 1.009 | 0.054 | 18.846 | *** | 0.713 |
| Q_35 | The doctors who treated me were highly knowledgeable. | <--- | Competency. | 1.003 | 0.052 | 19.354 | *** | 0.727 |
| Q_34 | The nurses were talking about me as if it was not there. | <--- | Competency. | 1.068 | 0.055 | 19.53 | *** | 0.741 |
| Q_33 | The hospital was discriminating in its treatment of patients. | <--- | Competency. | 0.995 | 0.052 | 19.144 | *** | 0.718 |
| Q_58 | The doctor who treated me was available during the holidays. | <--- | Available | 1 |  |  |  | 0.717 |
| Q_57 | The doctor who treated me was available all the time. | <--- | Available | 0.982 | 0.047 | 21.003 | *** | 0.736 |
| Q_56 | My physicians spent an appropriate amount of time with me. | <--- | Available | 0.917 | 0.047 | 19.646 | *** | 0.693 |

Table S3. Cont.

| Code | Question | Relation | HC Quality Subscales | Unstandardized Regression Weights | | | | Standardized Regression Weights |
| --- | --- | --- | --- | --- | --- | --- | --- | --- |
|  |  |  |  | **Estimate** | **S.E.** | **C.R.** | **P** | **Estimate** |
| Q_52 | I was kept informed about the results of tests and treatments. | <--- | Available | 1.021 | 0.049 | 20.711 | *** | 0.725 |
| Q_51 | My scheduled tests and procedures were performed on time. | <--- | Available | 0.966 | 0.045 | 21.509 | *** | 0.718 |
| Q_50 | I had to wait an unnecessarily long time to go to my room. | <--- | Available | 0.979 | 0.047 | 20.684 | *** | 0.724 |
| Q_49 | Usually, it took a long time to get the help I needed | <--- | Available | 0.951 | 0.048 | 19.868 | *** | 0.696 |
| Q_48 | I requested pain medicine; it took a long time before I got it. | <--- | Available | 0.918 | 0.046 | 19.923 | *** | 0.698 |
| Q_69 | The hospital conducts more medical tests than necessary. | <--- | Organize. | 1 |  |  |  | 0.698 |
| Q_71 | A lack of coordination between units or sections in the hospital | <--- | Organize. | 1.066 | 0.054 | 19.769 | *** | 0.721 |
| Q_72 | The administrative procedures were done correctly. | <--- | Organize. | 1.092 | 0.054 | 20.305 | *** | 0.742 |
| Q_73 | The admission process was quite organized. | <--- | Organize. | 1.095 | 0.055 | 19.786 | *** | 0.722 |
| Q_74 | The working hours for the cafeteria were convenient. | <--- | Organize. | 1.108 | 0.054 | 20.408 | *** | 0.747 |
| Q_75 | Visiting rules and regulations were established by the hospital. | <--- | Organize. | 1.012 | 0.053 | 19.215 | *** | 0.7 |
| Q_77 | The hospital staff is figuring out how to pay my hospital bills. | <--- | Organize. | 1.025 | 0.054 | 18.97 | *** | 0.69 |
| Q_78 | When I had to pay for the treatment, the procedures were simple. | <--- | Organize. | 1.031 | 0.054 | 19.176 | *** | 0.698 |
| Q_20 | The person who cleaned my room was friendly and courteous. | <--- | Empathy | 0.95 | 0.049 | 19.536 | *** | 0.726 |
| Q_19 | The nursing staff maintained and respected my privacy. | <--- | Empathy | 0.988 | 0.05 | 19.631 | *** | 0.721 |
| Q_18 | The admissions staff were friendly and helpful. | <--- | Empathy | 0.978 | 0.051 | 19.148 | *** | 0.71 |
| Q_17 | My family and visitors were treated with respect and dignity. | <--- | Empathy | 0.835 | 0.048 | 17.416 | *** | 0.646 |
| Q_16 | I was treated with respect and dignity. | <--- | Empathy | 0.99 | 0.053 | 18.76 | *** | 0.724 |
| Q_59 | There was a specialist present on all shifts. | <--- | Available | 0.953 | 0.047 | 20.415 | *** | 0.715 |
| Q_62 | I received written instructions when I was sent home. | <--- | Transition | 1 |  |  |  | 0.732 |
| Q_63 | Someone explained the purpose of the medicine I had | <--- | Transition | 1.074 | 0.05 | 21.298 | *** | 0.75 |
| Q_64 | Someone told me what danger signals to watch for after I went home. | <--- | Transition | 1.063 | 0.05 | 21.411 | *** | 0.746 |
| Q_65 | Someone told me about the medication's side effects. | <--- | Transition | 1.012 | 0.05 | 20.259 | *** | 0.716 |
| Q_66 | Someone told me when I could resume my usual activities. | <--- | Transition | 0.946 | 0.047 | 19.97 | *** | 0.697 |
| Q_67 | The doctors gave my family all the information they needed | <--- | Transition | 0.96 | 0.049 | 19.765 | *** | 0.69 |
| Q_68 | The nurses gave my family all the information they needed | <--- | Transition | 1.036 | 0.049 | 21.114 | *** | 0.736 |
| Q_60 | There was one particular doctor in charge of my care. | <--- | Available | 0.929 | 0.047 | 19.893 | *** | 0.696 |
| Q_61 | Then my family needed to confer with doctors, he was available. | <--- | Available | 0.949 | 0.047 | 20.283 | *** | 0.71 |
| Q_15 | I felt that the treatment of patients changed with time. | <--- | Empathy | 1 |  |  |  | 0.732 |
| Q_39 | The technical/administrative / support staff appeared professional. | <--- | Competency. | 1.033 | 0.054 | 19.255 | *** | 0.723 |
| Q_30 | One doctor said something, while another doctor said something else. | <--- | Competency. | 1 |  |  |  | 0.687 |

Note: ***indicates statistical significance.

Table S4: Standardized and Unstandardized Regression Weights for Health Care Quality, Patient Satisfaction, and Loyalty Model

| **Code** | **Question** | **Relation** | **HC Quality Subscales** | **Unstandardized Regression Weights** | | | | **Standardized Regression Weights** |
| --- | --- | --- | --- | --- | --- | --- | --- | --- |
|  |  |  |  | **Estimate** | **S.E.** | **C.R.** | **P** | **Estimate** |
| **Q_1** | The cleanliness throughout the hospital | <--- | Tangible | 1 |  |  |  | 0.707 |
| **Q_2** | The cabins/wards were clean. | <--- | Tangible | 0.983 | 0.049 | 20.173 | *** | 0.716 |
| **Q_3** | The doctors, nurses, and staff were neatly dressed. | <--- | Tangible | 1.093 | 0.053 | 20.662 | *** | 0.734 |
| **Q_5** | The hospital has modern-looking equipment. | <--- | Tangible | 0.978 | 0.049 | 19.869 | *** | 0.705 |
| **Q_6** | The hospital used state-of-the-art medical equipment. | <--- | Tangible | 0.98 | 0.049 | 19.984 | *** | 0.709 |
| **Q_8** | The physical facilities were visually appealing. | <--- | Tangible | 1.011 | 0.049 | 20.558 | *** | 0.73 |
| **Q_9** | The toilets were clean. | <--- | Tangible | 0.977 | 0.048 | 20.366 | *** | 0.723 |
| **Q_10** | Adequate parking was available. | <--- | Tangible | 0.995 | 0.05 | 20.077 | *** | 0.713 |
| **Q_12** | I found the taste of my food to be very good. | <--- | Tangible | 0.93 | 0.05 | 18.462 | *** | 0.654 |
| **Q_13** | Specially assigned rooms were available. | <--- | Tangible | 1.014 | 0.052 | 19.695 | *** | 0.699 |
| **Q_40** | It was easy for me to find someone to talk with. | <--- | Information | 1 |  |  |  | 0.748 |
| **Q_41** | My family had enough opportunity to talk to my doctors. | <--- | Information | 0.991 | 0.048 | 20.796 | *** | 0.73 |
| **Q_42** | My family had enough opportunity to talk to the nurses. | <--- | Information | 1 | 0.048 | 21.017 | *** | 0.737 |
| **Q_44** | The hospital informed me about the medical options. | <--- | Information | 0.951 | 0.047 | 20.368 | *** | 0.715 |
| **Q_45** | I got enough information about my medical condition. | <--- | Information | 0.925 | 0.046 | 20.072 | *** | 0.705 |
| **Q_28** | I got answers that I could understand from doctors. | <--- | Response | 1 |  |  |  | 0.716 |
| **Q_27** | I got answers that I could understand from the Nurse. | <--- | Response | 1.091 | 0.05 | 21.625 | *** | 0.759 |
| **Q_26** | The nursing staff explained the treatment simply. | <--- | Response | 1.018 | 0.049 | 20.733 | *** | 0.728 |
| **Q_25** | Someone explained to me the reasons why I had to wait. | <--- | Response | 0.991 | 0.047 | 20.871 | *** | 0.733 |
| **Q_24** | I had said enough about the medical treatment. | <--- | Response | 1.081 | 0.049 | 22.044 | *** | 0.774 |
| **Q_23** | The hospital involved me in the decision about my care. | <--- | Response | 1.088 | 0.05 | 21.967 | *** | 0.771 |
| **Q_22** | When I had some anxiety or fear, the nurse discussed it. | <--- | Response | 1.035 | 0.05 | 20.818 | *** | 0.731 |
| **Q_21** | When I had some anxiety or fear, the doctor discussed it. | <--- | Response | 1.026 | 0.049 | 21.114 | *** | 0.741 |
| **Q_38** | The hospital did all it could to help control my pain. | <--- | Competent | 1.022 | 0.053 | 19.13 | *** | 0.714 |
| **Q_37** | I had full confidence and trust in the nurse treating me. | <--- | Competent | 1.026 | 0.053 | 19.413 | *** | 0.725 |
| **Q_35** | The doctors who treated me were highly knowledgeable. | <--- | Competent | 0.996 | 0.052 | 19.334 | *** | 0.722 |
| **Q_34** | The nurses were talking about me as if it was not there. | <--- | Competent | 1.089 | 0.054 | 20.123 | *** | 0.755 |
| **Q_33** | The hospital was discriminating in its treatment of patients. | <--- | Competent | 0.993 | 0.052 | 19.202 | *** | 0.717 |
| **Q_58** | The doctor who treated me was available during the holidays. | <--- | Available | 1 |  |  |  | 0.714 |
| **Q_57** | The doctor who treated me was available all the time. | <--- | Available | 0.988 | 0.047 | 20.838 | *** | 0.737 |
| **Q_56** | My physicians spent an appropriate amount of time with me. | <--- | Available | 0.918 | 0.047 | 19.564 | *** | 0.691 |
| **Q_52** | I was kept informed about the results of tests and treatments. | <--- | Available | 1.027 | 0.05 | 20.551 | *** | 0.727 |
| **Q_51** | My scheduled tests and procedures were performed on time. | <--- | Available | 0.97 | 0.045 | 21.396 | *** | 0.718 |
| **Q_50** | I had to wait an unnecessarily long time to go to my room. | <--- | Available | 0.989 | 0.049 | 20.085 | *** | 0.729 |
| **Q_49** | Usually, it took a long time to get the help I needed | <--- | Available | 0.945 | 0.046 | 20.579 | *** | 0.688 |
| **Q_48** | I requested pain medicine; it took a long time before I got it. | <--- | Available | 0.923 | 0.047 | 19.771 | *** | 0.698 |
| **Q_69** | The hospital conducts more medical tests than necessary. | <--- | Organize | 1 |  |  |  | 0.701 |
| **Q_71** | A lack of coordination between units or sections in the hospital | <--- | Organize | 1.062 | 0.053 | 19.884 | *** | 0.722 |
| **Q_72** | The administrative procedures were done correctly. | <--- | Organize | 1.087 | 0.053 | 20.405 | *** | 0.742 |
| **Q_73** | The admission process was quite organized. | <--- | Organize | 1.09 | 0.055 | 19.878 | *** | 0.722 |
| **Q_74** | The working hours for the cafeteria were convenient. | <--- | Organize | 1.101 | 0.054 | 20.465 | *** | 0.745 |
| **Q_75** | Visiting rules and regulations were established by the hospital. | <--- | Organize | 1.009 | 0.052 | 19.324 | *** | 0.701 |
| **Q_77** | The hospital staff is figuring out how to pay my hospital bills. | <--- | Organize | 1.021 | 0.054 | 19.056 | *** | 0.69 |
| **Q_78** | When I had to pay for the treatment, the procedures were simple. | <--- | Organize | 1.025 | 0.053 | 19.224 | *** | 0.697 |
| **Q_20** | The person who cleaned my room was friendly and courteous. | <--- | Empathy | 0.978 | 0.049 | 19.808 | *** | 0.733 |
| **Q_19** | The nursing staff maintained and respected my privacy. | <--- | Empathy | 1.006 | 0.051 | 19.672 | *** | 0.721 |
| **Q_18** | The admissions staff were friendly and helpful. | <--- | Empathy | 0.997 | 0.052 | 19.147 | *** | 0.711 |
| **Q_17** | My family and visitors were treated with respect and dignity. | <--- | Empathy | 0.861 | 0.049 | 17.671 | *** | 0.654 |
| **Q_16** | I was treated with respect and dignity. | <--- | Empathy | 0.986 | 0.052 | 19.066 | *** | 0.708 |
| **Q_59** | There was a specialist present on all shifts. | <--- | Available | 0.955 | 0.047 | 20.196 | *** | 0.714 |
| **Q_62** | I received written instructions when I was sent home. | <--- | Transition | 1 |  |  |  | 0.73 |
| **Q_63** | Someone explained the purpose of the medicine I had | <--- | Transition | 1.071 | 0.05 | 21.203 | *** | 0.746 |
| **Q_64** | Someone told me what danger signals to watch after I went home. | <--- | Transition | 1.08 | 0.05 | 21.492 | *** | 0.756 |
| **Q_65** | Someone told me about the medication's side effects. | <--- | Transition | 1.013 | 0.05 | 20.272 | *** | 0.714 |
| **Q_66** | Someone told me when I could resume my usual activities. | <--- | Transition | 0.948 | 0.047 | 19.974 | *** | 0.696 |
| **Q_67** | The doctors gave my family all the information they needed | <--- | Transition | 0.979 | 0.049 | 19.877 | *** | 0.701 |
| **Q_68** | The nurses gave my family all the information they needed | <--- | Transition | 1.035 | 0.049 | 21.045 | *** | 0.733 |
| **Q_60** | There was one particular doctor in charge of my care. | <--- | Available | 0.934 | 0.047 | 19.74 | *** | 0.697 |
| **Q_61** | Then my family needed to confer with doctors, he was available. | <--- | Available | 0.953 | 0.047 | 20.107 | *** | 0.71 |
| **Q_15** | I felt that the treatment of patients changed with time. | <--- | Empathy | 1 |  |  |  | 0.719 |
| **Q_39** | The technical/administrative / support staff appeared professional. | <--- | Competent | 1.027 | 0.053 | 19.248 | *** | 0.718 |
| **Q_30** | One doctor said something, while another doctor said something else. | <--- | Competent | 1 |  |  |  | 0.687 |
| **Q_84** | Overall, I was satisfied with the care I received at this hospital. | <--- | Satisfaction | 1 |  |  |  | 0.697 |
| **Q_83** | I received courteous and professional care while in the hospital. | <--- | Satisfaction | 1.1 | 0.055 | 20.113 | *** | 0.727 |
| **Q_82** | I am satisfied with the care provided by this hospital. | <--- | Satisfaction | 1.136 | 0.055 | 20.531 | *** | 0.744 |
| **Q_81** | This hospital is considered to be a good place for care. | <--- | Satisfaction | 1.048 | 0.05 | 20.965 | *** | 0.724 |
| **Q_80** | I am satisfied with the treatment and outcome of care. | <--- | Satisfaction | 1.112 | 0.054 | 20.688 | *** | 0.753 |
| **Q_85** | I would be willing to return to this hospital in the future if needed. | <--- | loyalty | 1.028 | 0.05 | 20.386 | *** | 0.725 |
| **Q_86** | I feel comfortable recommending the hospital to my friend. | <--- | loyalty | 1.069 | 0.051 | 21.122 | *** | 0.752 |
| **Q_87** | I would recommend the hospital to my family. | <--- | loyalty | 1 |  |  |  | 0.703 |

Note: ***indicates statistical significance.

Table S5: Standardized and Unstandardized regression weights for the measurement model examining pooled Health Care quality components, patient satisfaction, and loyalty.

| Code | Question | Relation | HC Quality Subscales | Unstandardized Regression Weights | | | | Standardized Regression Weights |
| --- | --- | --- | --- | --- | --- | --- | --- | --- |
|  |  |  |  | **Estimate** | **S.E.** | **C.R.** | **P** | **Estimate** |
| Tangible | Tangibles and Physical attributes | <--- | HC Quality | 1 |  |  |  | 0.412 |
| Empathy | Empathy and Courtesy | <--- | HC Quality | 1.273 | 0.122 | 10.449 | *** | 0.55 |
| Response | Responsiveness and patient involvement | <--- | HC Quality | 0.771 | 0.102 | 7.587 | *** | 0.319 |
| Competency | Competency, Confidence, and Trust | <--- | HC Quality | 0.979 | 0.108 | 9.089 | *** | 0.421 |
| Information | Communication and Information | <--- | HC Quality | 1.658 | 0.146 | 11.352 | *** | 0.676 |
| Availability | Availability, Accessibility, and Timelines | <--- | HC Quality | 0.926 | 0.104 | 8.865 | *** | 0.404 |
| Transition | Transition of care | <--- | HC Quality | 1.184 | 0.118 | 10.008 | *** | 0.503 |
| Organization | Organization rules, processes, and payments | <--- | HC Quality | 0.806 | 0.101 | 8.009 | *** | 0.345 |
| Q_80 | I am satisfied with the outcome of care. | <--- | Satisfaction | 1 |  |  |  | 0.743 |
| Q_81 | This hospital is a good place for care. | <--- | Satisfaction | 0.947 | 0.045 | 21.265 | *** | 0.719 |
| Q_82 | I am satisfied with the care provided | <--- | Satisfaction | 1.033 | 0.047 | 22.031 | *** | 0.743 |
| Q_83 | I received courteous and professional care. | <--- | Satisfaction | 1.003 | 0.046 | 21.586 | *** | 0.729 |
| Q_84 | Overall, I was satisfied with the care. | <--- | Satisfaction | 0.919 | 0.044 | 20.8 | *** | 0.704 |
| Q_85 | I would be willing to return to this hospital. | <--- | Loyalty | 1 |  |  |  | 0.725 |
| Q_86 | I will recommend the hospital to my friend. | <--- | Loyalty | 1.036 | 0.048 | 21.705 | *** | 0.75 |
| Q_87 | I would recommend the hospital to my family. | <--- | Loyalty | 0.973 | 0.048 | 20.325 | *** | 0.703 |

Note: ***indicates statistical significance
