## Supplementary material for "Structural Equation Modelling of Healthcare Quality, Patient Satisfaction, and Patient Loyalty in Health Insurance Hospitals in Alexandria, Egypt": Suppl figures

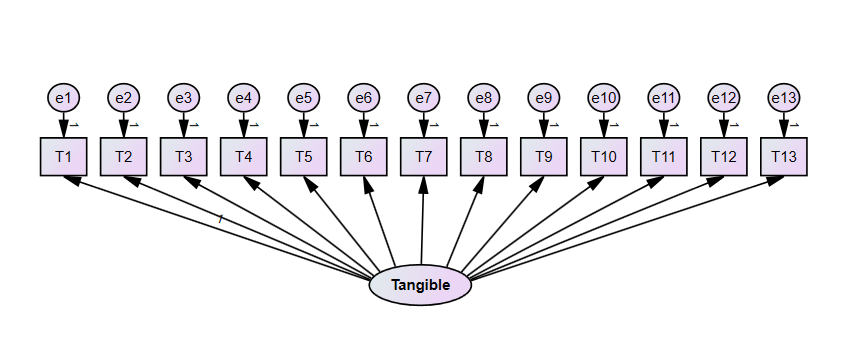

Figure S1: Tangible, ancillary services and facilities theoretical model

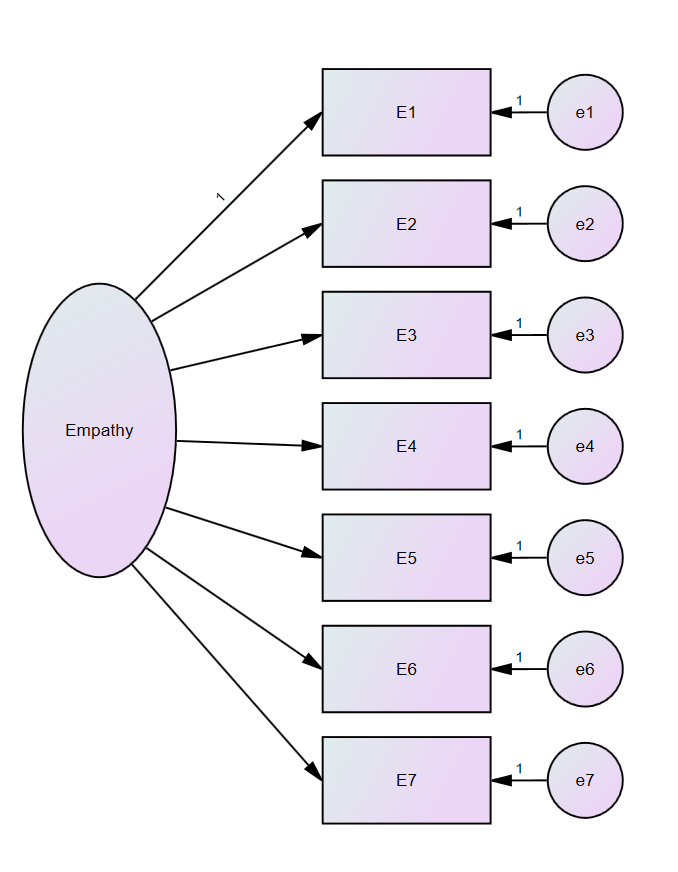

Figure S2: Courtesy and Empathy theoretical model

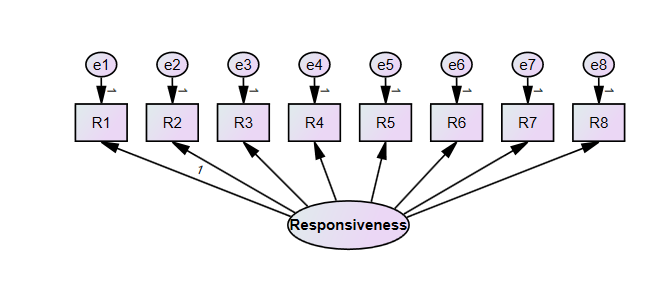

Figure S3: Responsiveness and physiological aspects and the Patient Involvement Theoretical Model

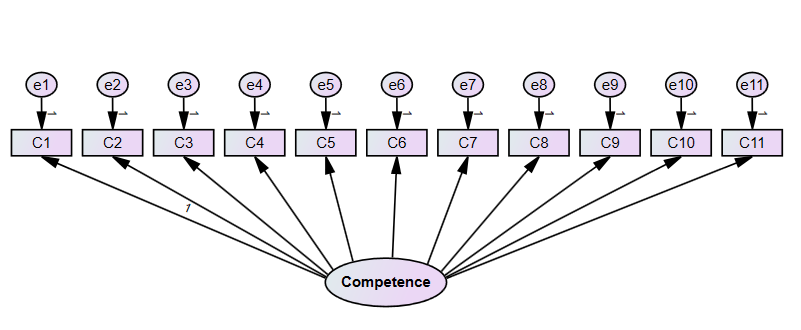

Figure S4: Competency, confidence, Fairness, and trust theoretical model

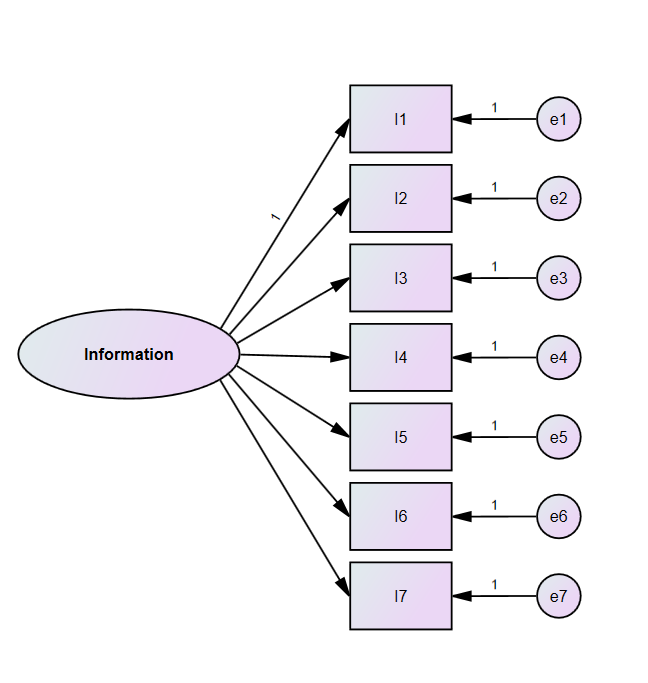

Figure S5: Information and Communication Theoretical Model

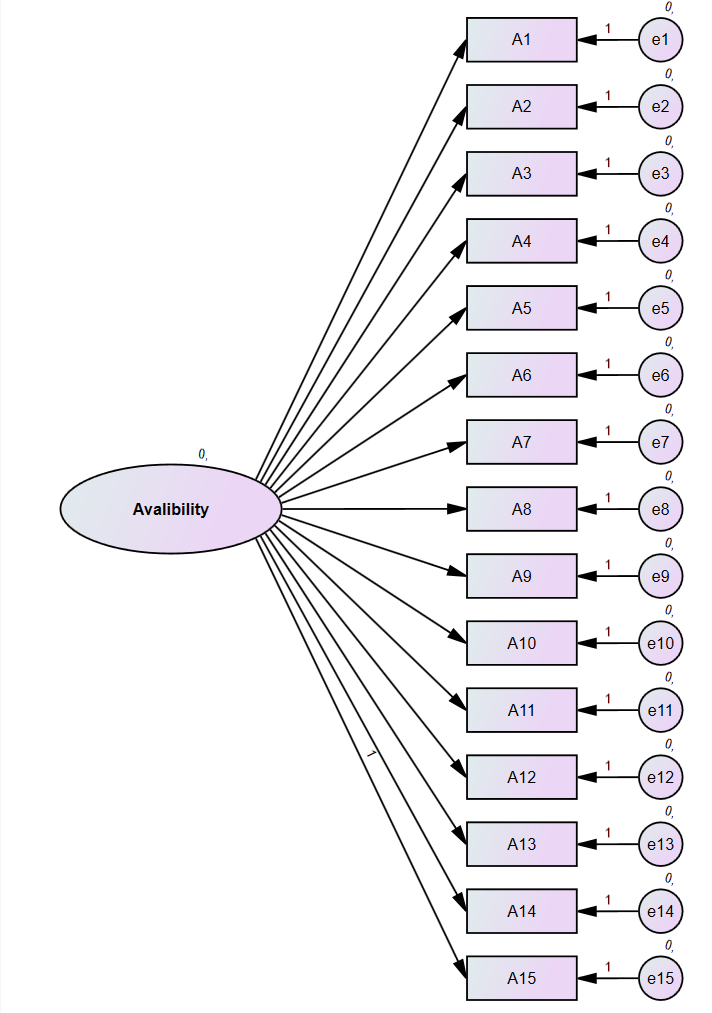

Figure S6: Availability, accessibility, and Timeliness of services theoretical model

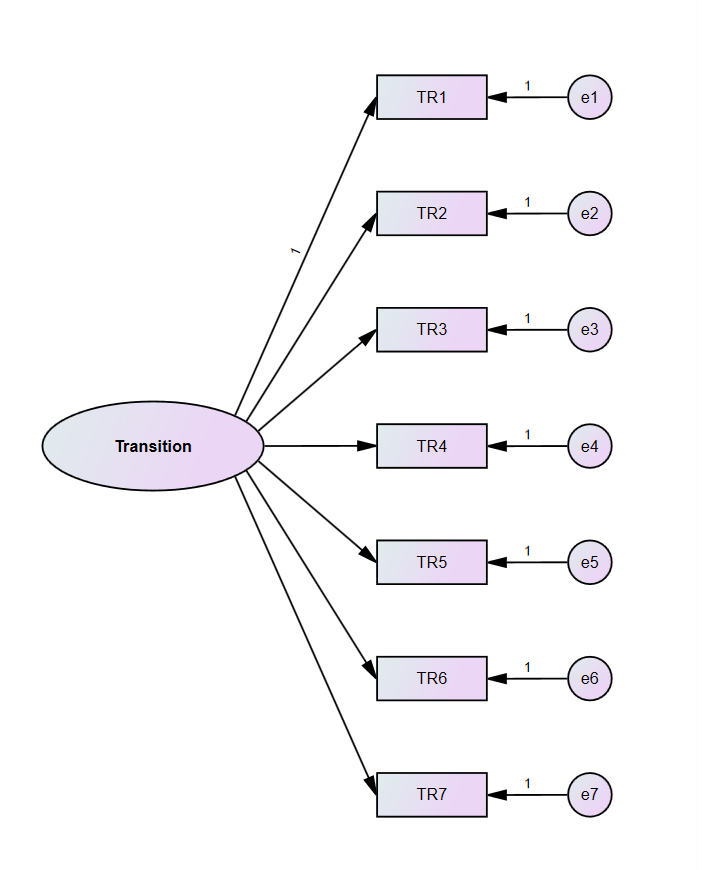

Figure S7: Transition to home theoretical model

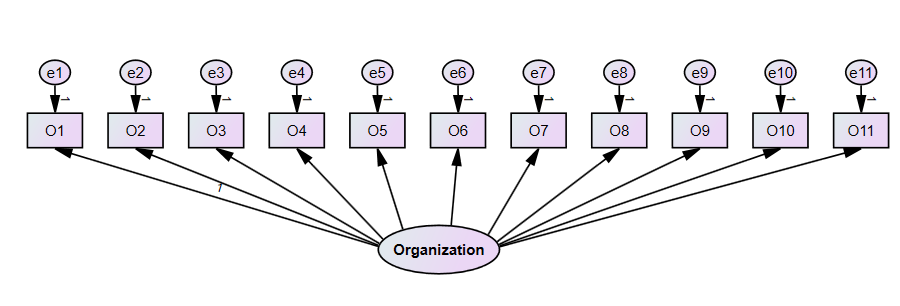

Figure S8: Organization Management, Regulations, and Payment Theoretical Model

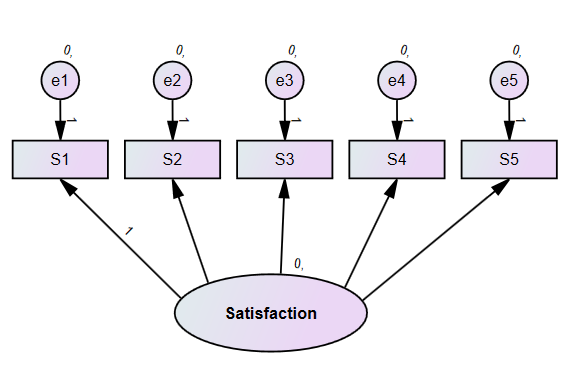

Figure S9: Patient Satisfaction theoretical model

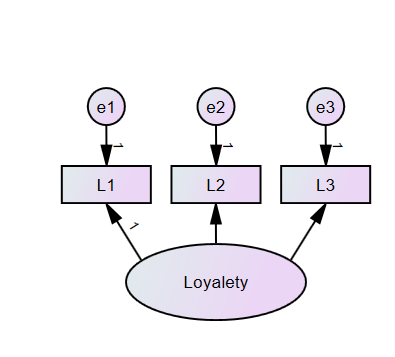

Figure S10: Patient Loyalty Theoretical Model

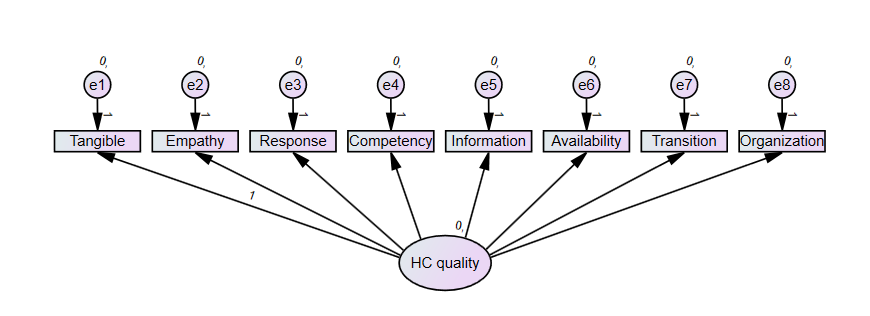

Figure S11: Healthcare Quality Component Measurement Model

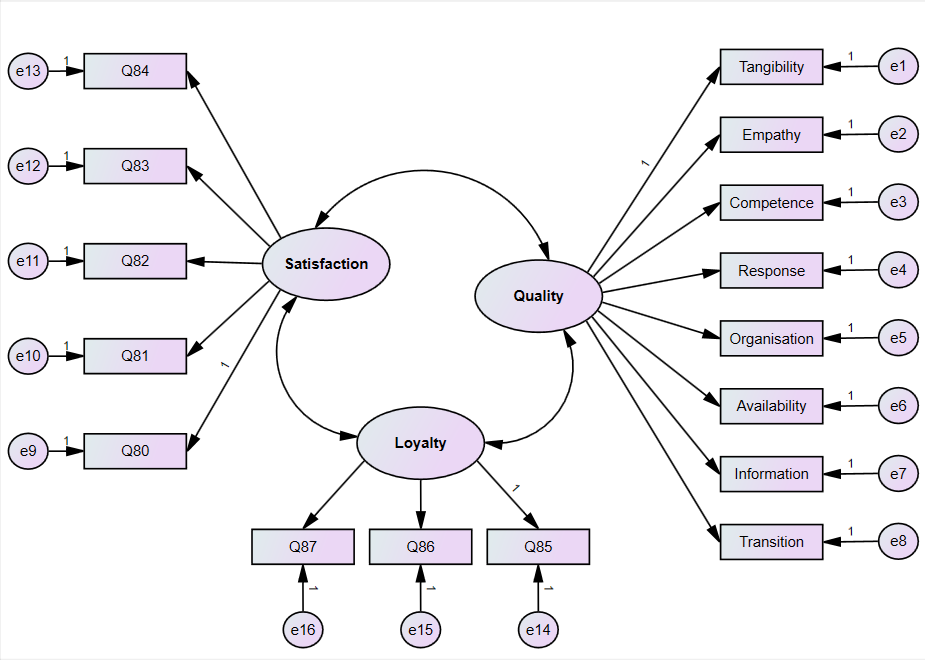

Figure S12:	Study model of causal relationships between healthcare Quality, Patient Satisfaction, and Patient Loyalty

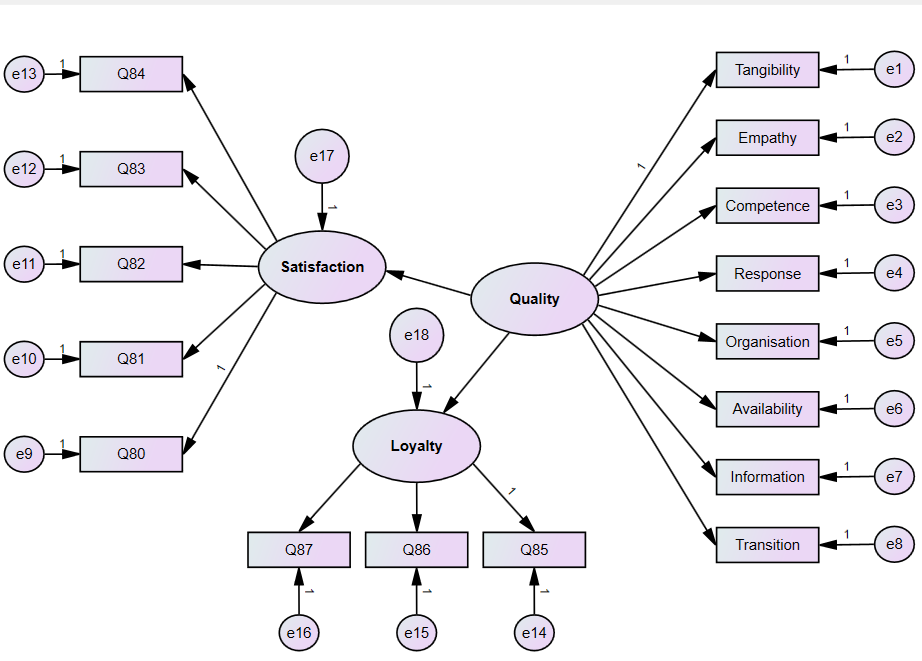

Figure S13: Study Model for the equal effect of the healthcare Quality on Patient Satisfaction and Loyalty

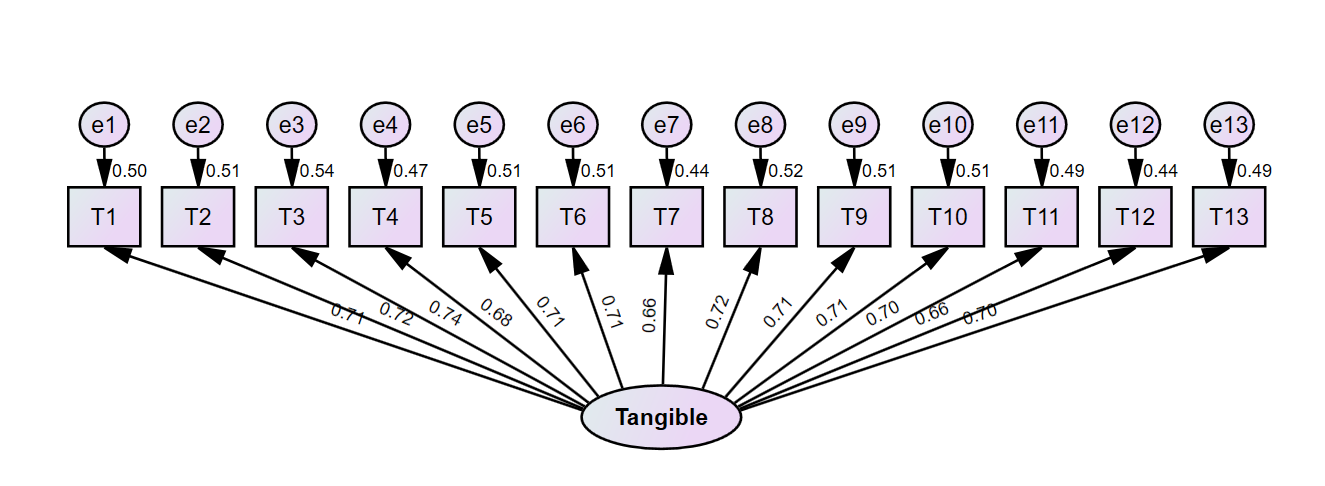

Figure S14: Measurement model of the Tangible, facility, and ancillary services attributes

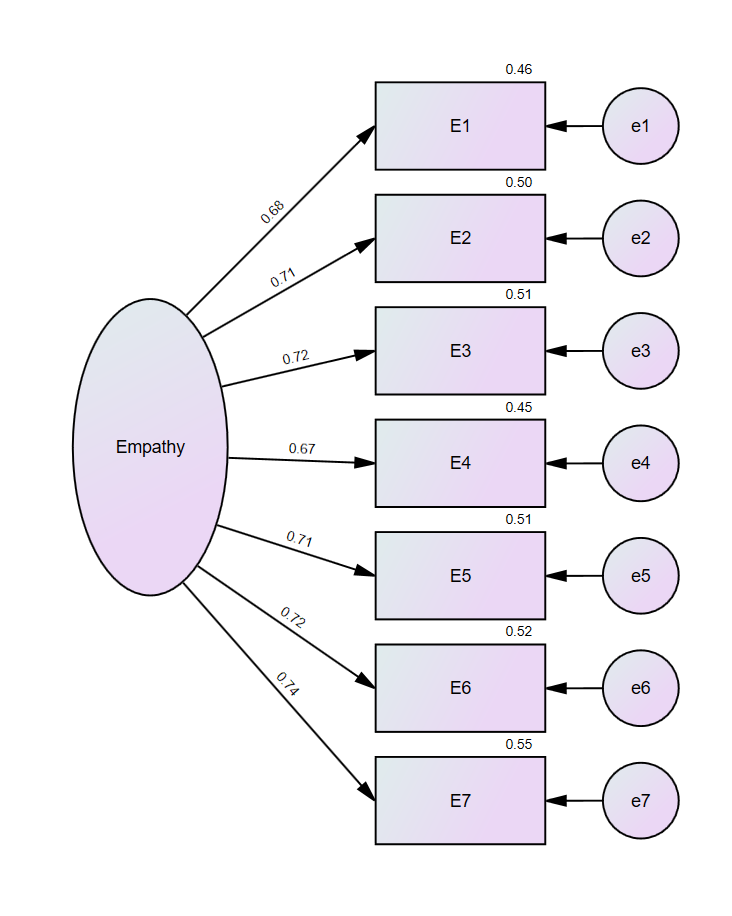

Figure S15: Measurement model of Empathy and personal attention to the patient attributes

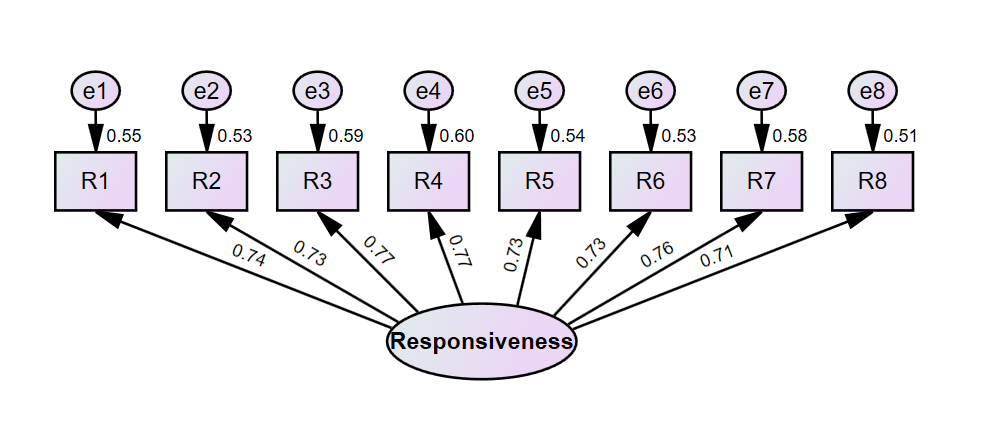

Figure S16: Measurement model of Responsiveness and patient involvement attributes

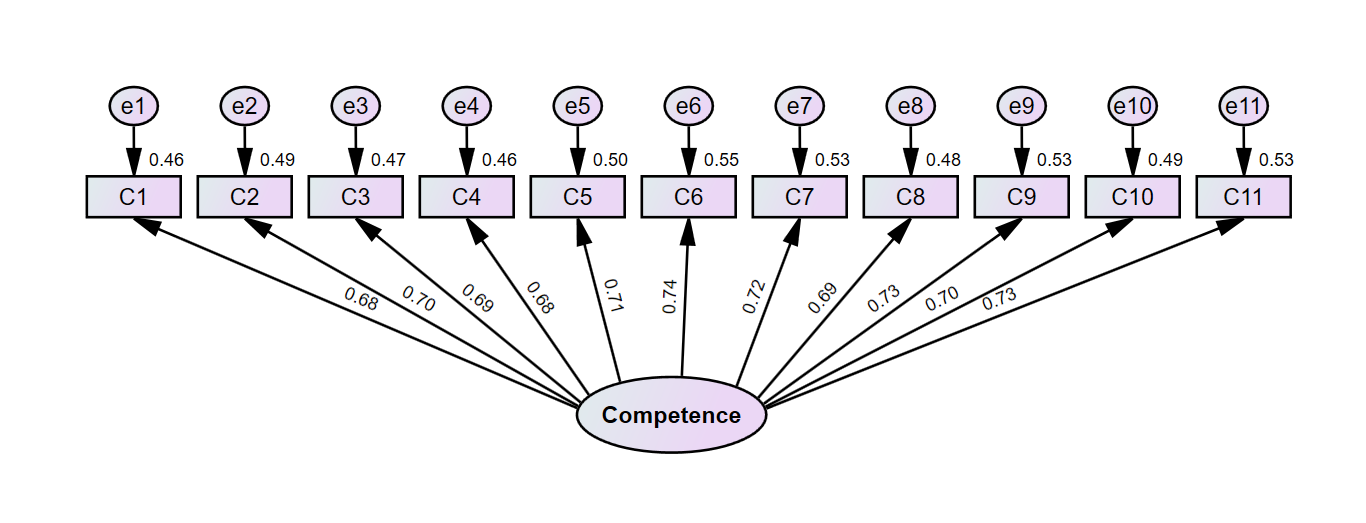

Figure S17: Measurement model of Competence, Knowledge, Confidence, and trust attributes

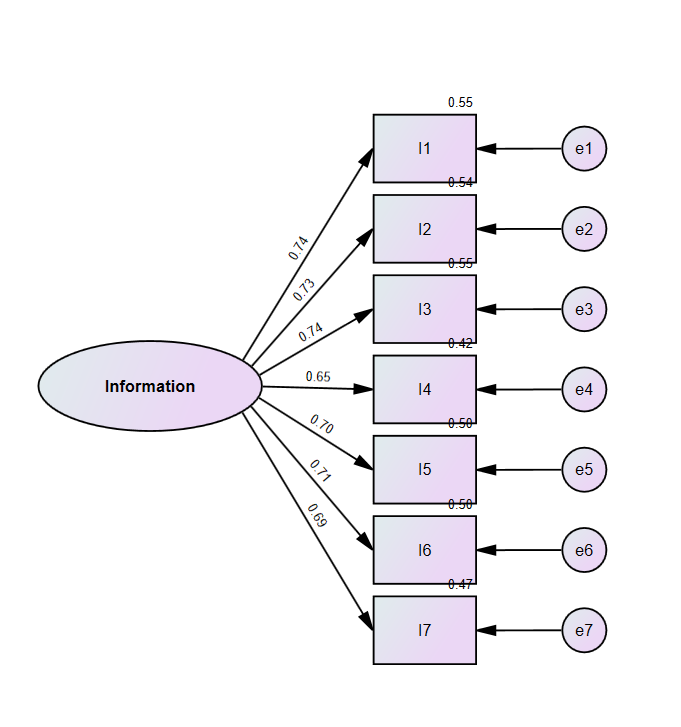

Figure S18: Measurement model of the Information and Communication attributes

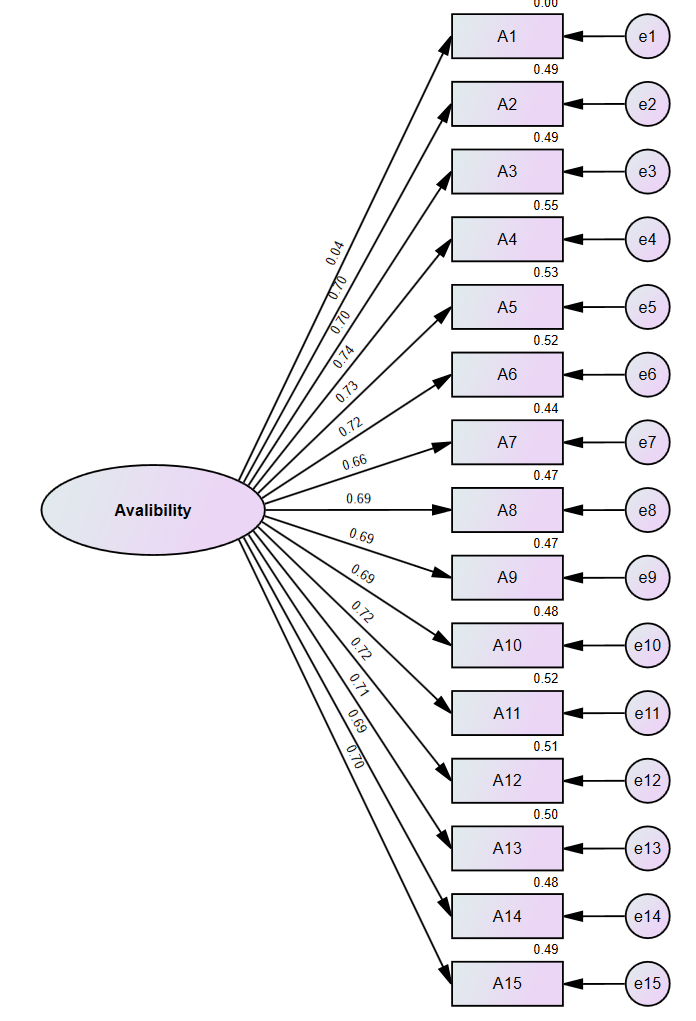

Figure S19: Measurement model of Availability, Accessibility, and waiting time attributes

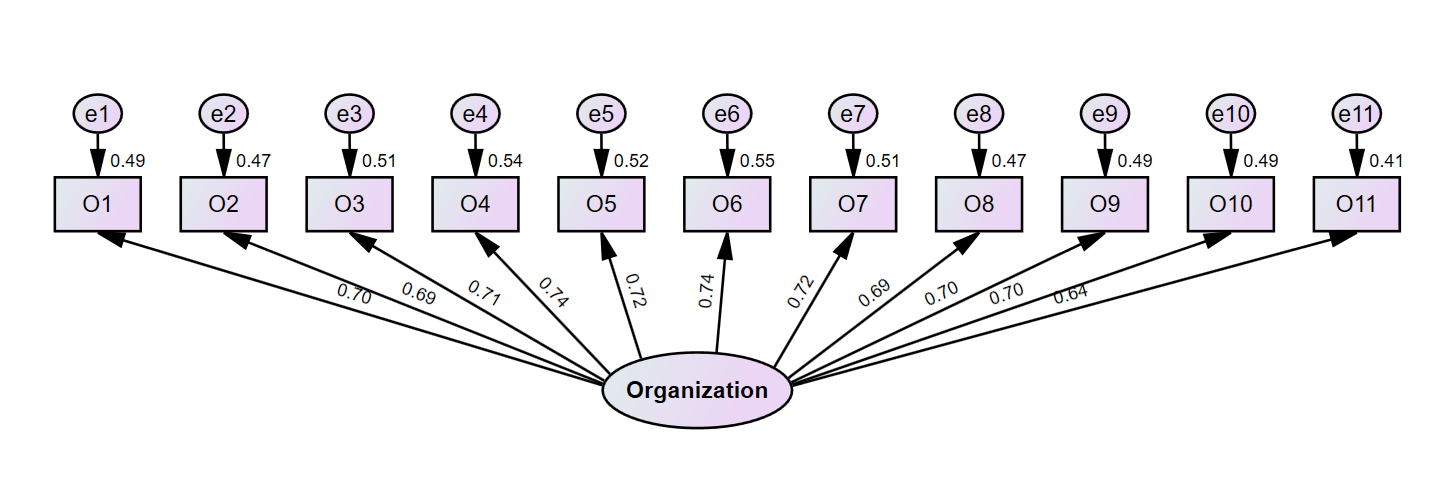

Figure S20: Measurement model of the Hospital Procedures, Organization, Administration, and Payment attributes

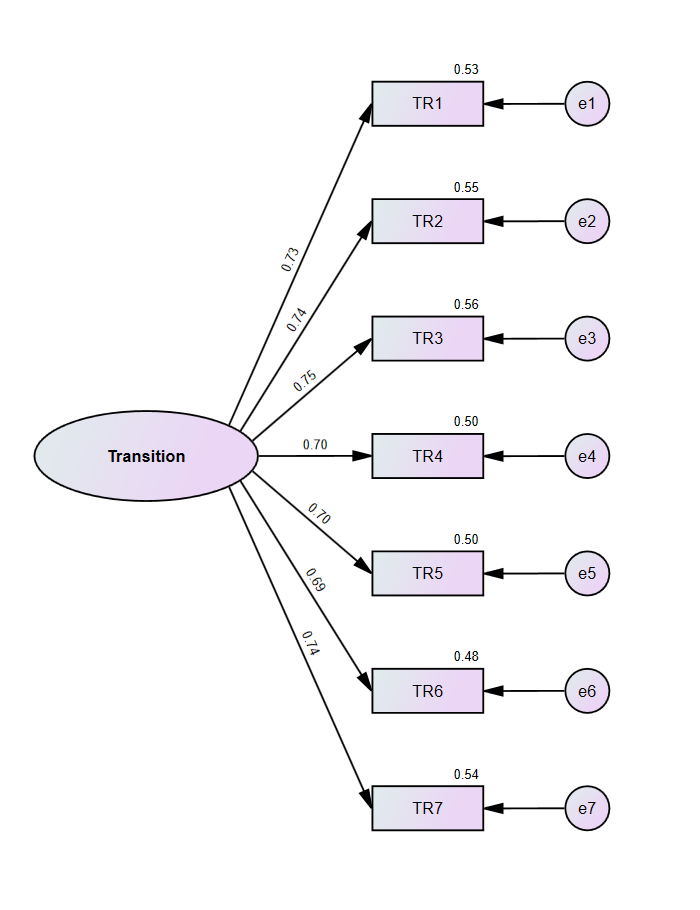

Figure S21: Measurement model of the Transition to home attributes

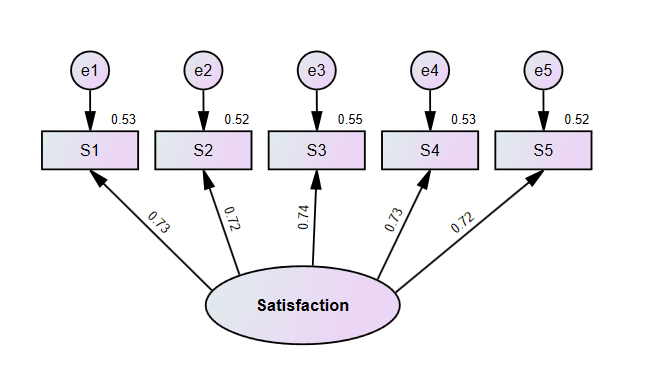

Figure S22: Measurement model of the Patient Satisfaction Attributes

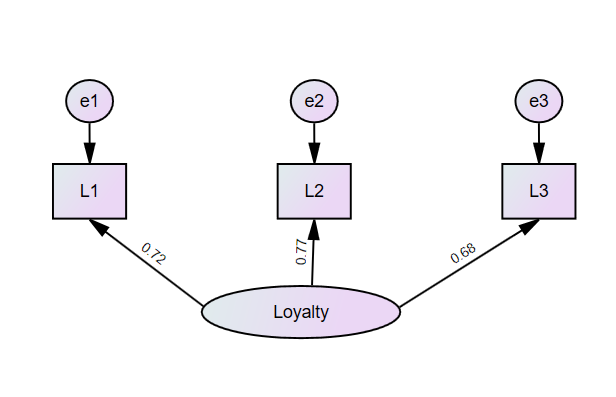

Figure S23: Measurement model of the Patient Loyalty Attributes

| 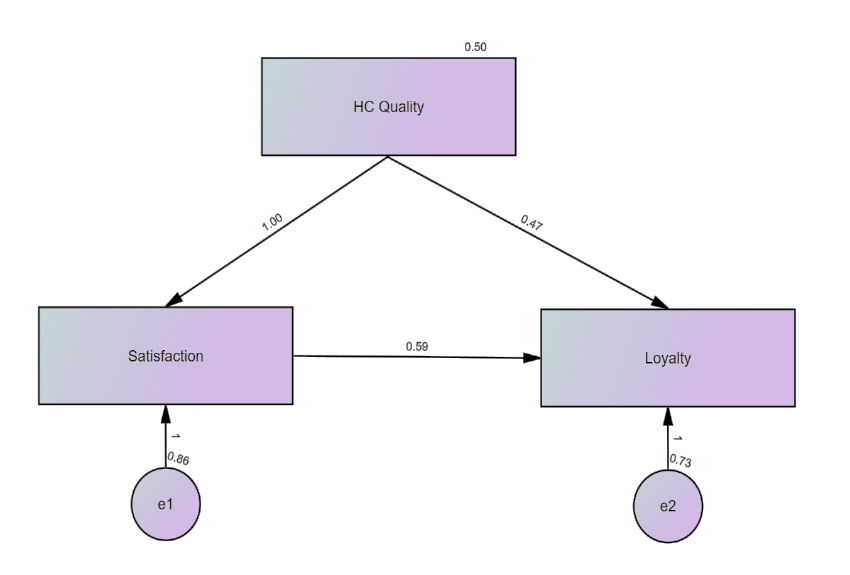 | 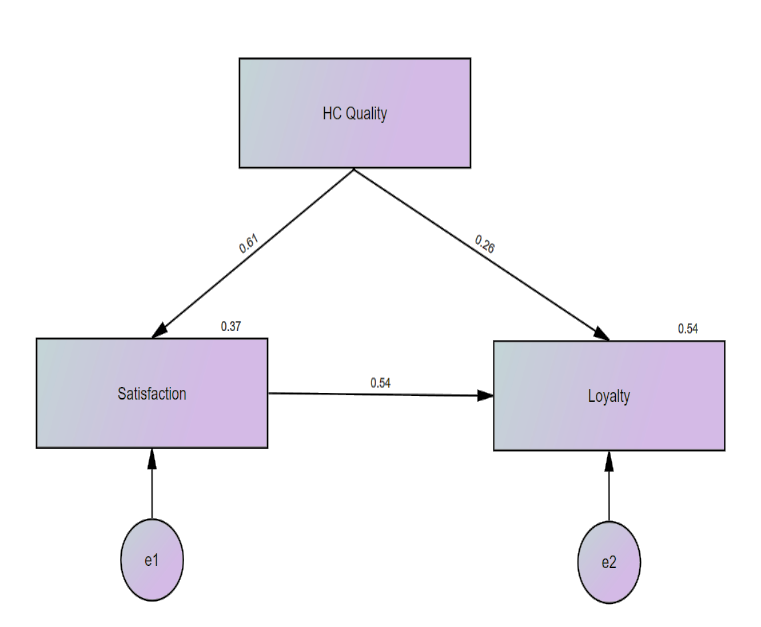 |
| --- | --- |

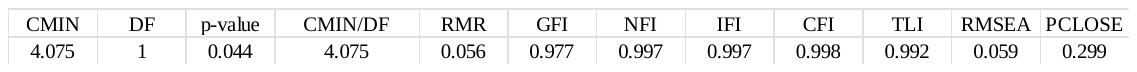

Figure S24:	Unstandardized and Standardized Path Analysis of the HQ Quality, Patient Satisfaction, and Patient Loyalty

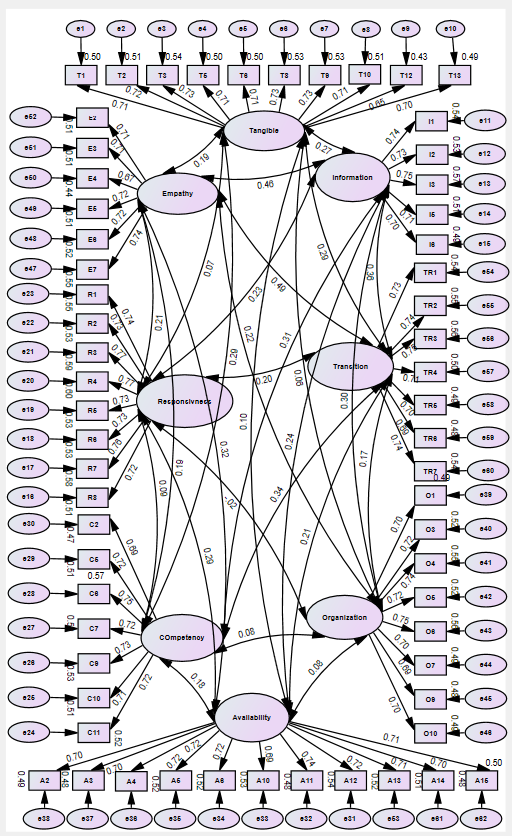

| Index | Value |
| --- | --- |
| χ² | 3621.338 |
| DF | 2335 |
| P-value | 0.000 |
| CMIN/DF | 1.979 |
| RMR | 0.080 |
| GFI | 0.875 |
| NFI | 0.875 |
| IFI | 0.928 |
| CFI | 0.928 |
| TLI | 0.925 |
| RMSEA | 0.033 |
| PCLOSE | 1.000 |

Figure S25 : Initial Measurement Model of Health Care Quality Construct

| Index | Value |
| --- | --- |
| χ² | 3317.961 |
| DF | 1793 |
| P-value | 0.000 |
| CMIN/DF | 1.851 |
| RMR | 0.057 |
| GFI | 0.896 |
| NFI | 0.888 |
| TLI | 0.940 |
| IFI | 0.944 |
| CFI | 0.943 |
| RMSEA | 0.031 |
| PCLOSE | 1.000 |

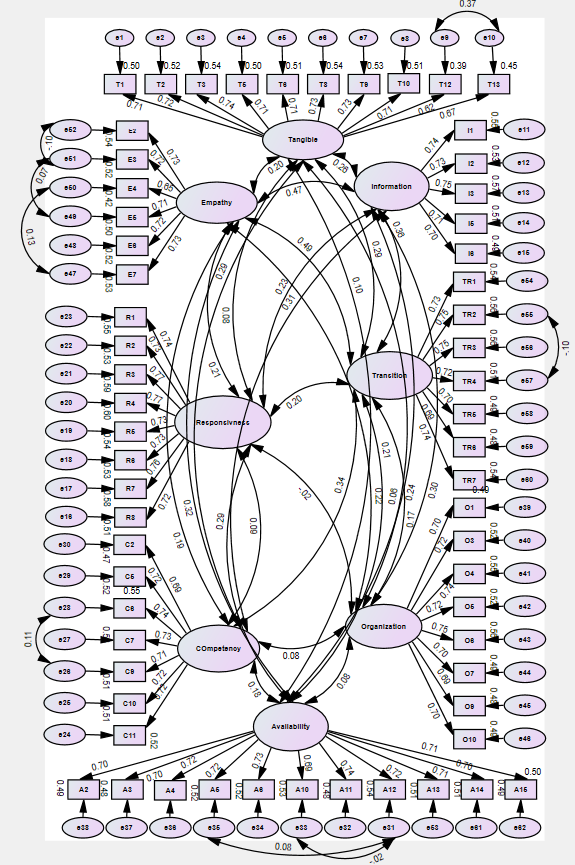

Figure S26: Modified Healthcare Quality Measurement Model

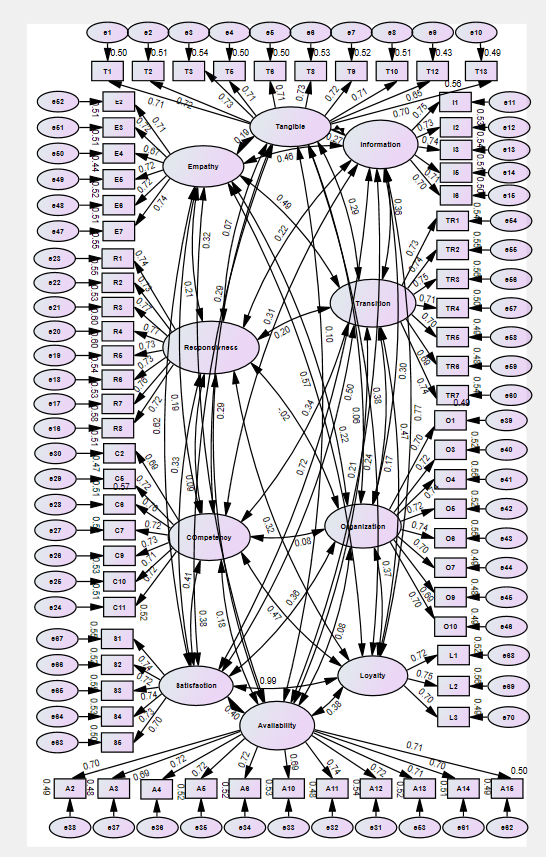

| Index | Value |
| --- | --- |
| χ² | 266.676 |
| DF | 101 |
| P-value | 0.000 |
| CMIN/DF | 2.640 |
| RMR | 0.048 |
| GFI | 0.951 |
| NFI | 0.948 |
| IFI | 0.967 |
| CFI | 0.967 |
| TLI | 0.967 |
| RMSEA | 0.043 |
| PCLOSE | 0.967 |

Figure S27: Health Care Quality, Patient Satisfaction, and Patient Loyalty Measurement Model

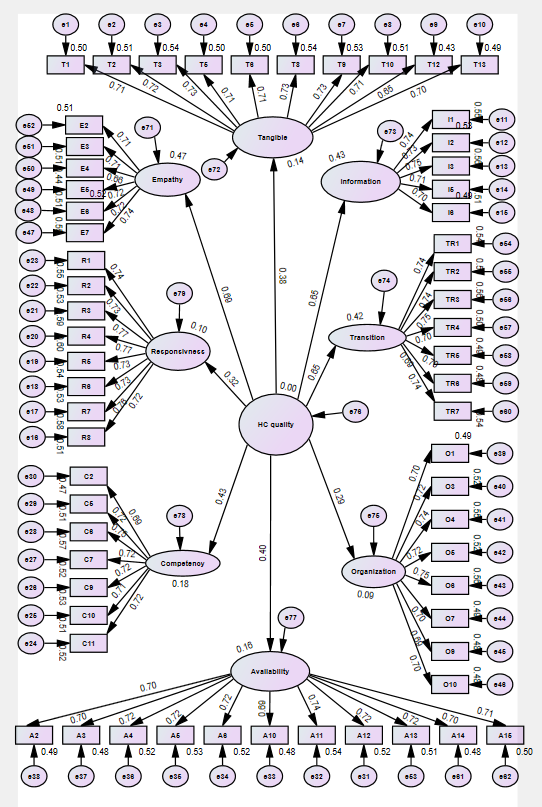

Figure S28: The relationship between health care quality and the observed variables

| Index | Value |
| --- | --- |
| χ² | 4423.566 |
| DF | 2300.000 |
| P-value | 0.000 |
| CMIN/DF | 1.923 |
| RMR | 0.057 |
| GFI | 0.880 |
| NFI | 0.870 |
| IFI | 0.933 |
| CFI | 0.933 |
| TLI | 0.923 |
| RMSEA | 0.032 |
| PCLOSE | 1.000 |

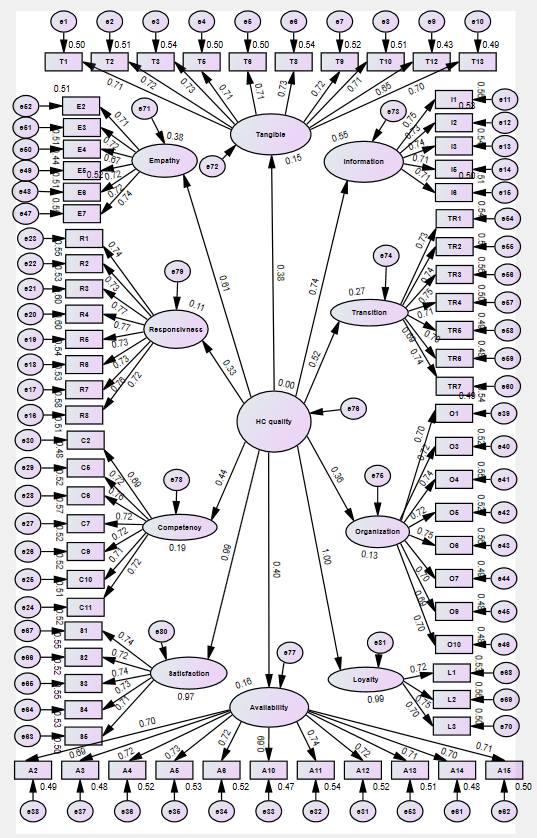

Figure S29: The relationship between healthcare quality observed variables, patient satisfaction, and loyalty

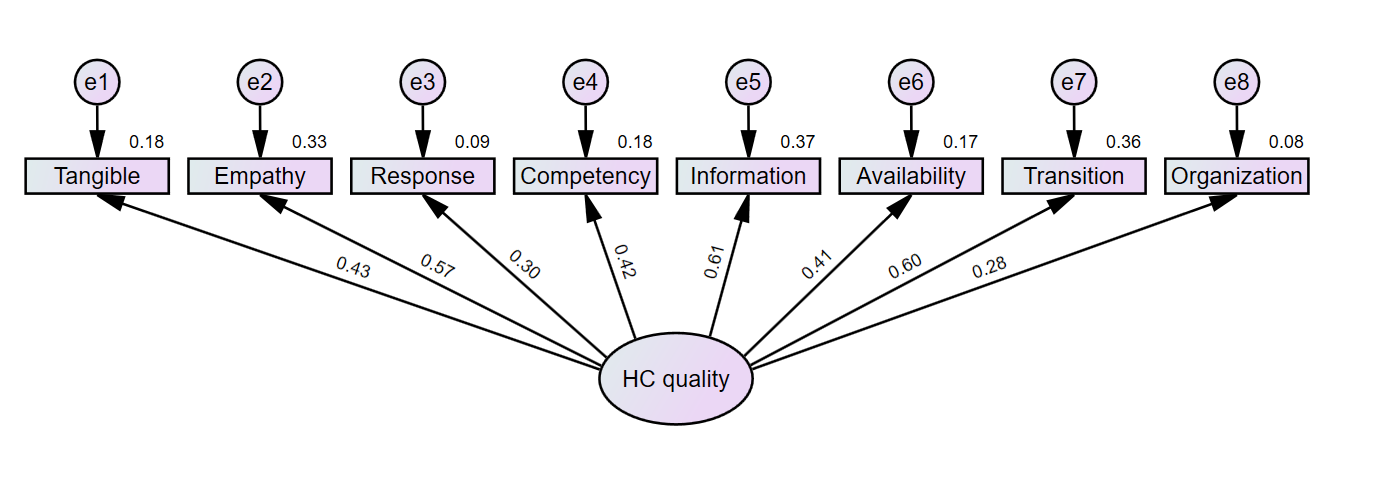

Figure S30: The Health Care Quality construct

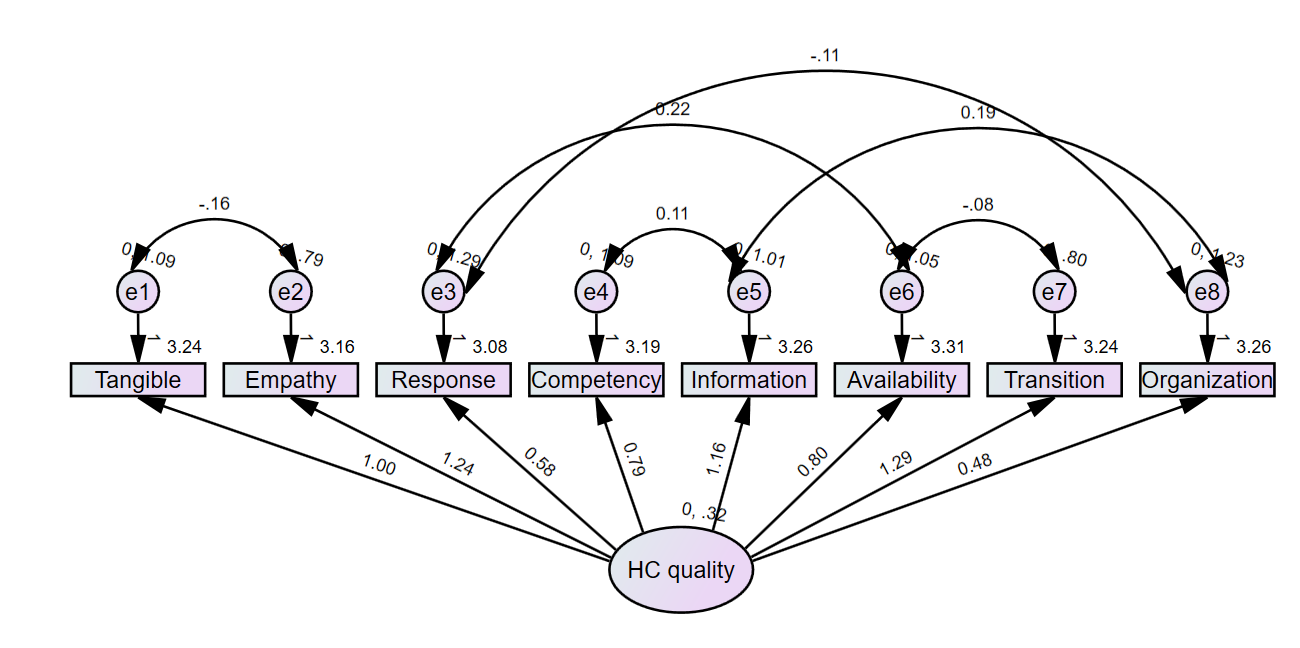

Figure S31: The Modified Health Care Quality construct

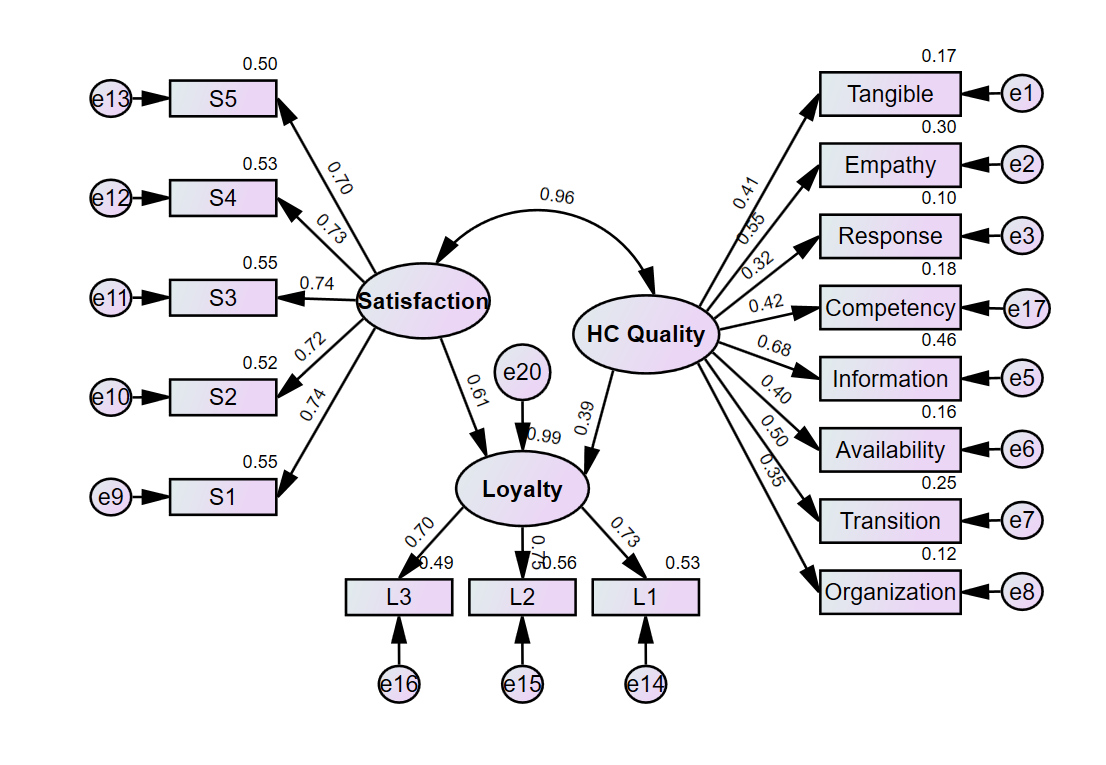

Figure S32: Theoretical model of the two paths relationships between healthcare quality, patient satisfaction, and patient loyalty
